# Developing an intervention to maintain biweekly asymptomatic SARS-CoV-2 testing amongst English care home staff: An integrative approach

**DOI:** 10.1101/2025.06.10.25329242

**Authors:** Paul Flowers, Ruth Leiser, Maria Krutikov, Natalie Adams, Hadjer Nacer-Laidi, Julie McLeod, Catherine Henderson, Borscha Azmi, Dorina Cadar, Lara Goscé, Oliver Stirrup, James Blackstone, Laura Shallcross

## Abstract

**Purpose:** To describe the development of an intervention (‘*Test to Care’*) maintaining biweekly asymptomatic SARS-CoV-2 testing amongst care home staff.

**Methods:** Diverse inputs were sequentially integrated: *1*) A behaviour change wheel (BCW) analysis of the results from a systematic review of barriers and facilitators to staff testing for SARS-CoV-2 from 14 international studies to generate initial intervention content ideas; *2*) A series of eight stakeholder events with UK care home staff and policy makers (N=∼70) to iteratively operationalise emerging intervention content; *3*) Confirmatory thematic analysis of barriers and facilitators to biweekly asymptomatic SARS-CoV-2 testing from four focus groups (N=15) to check temporal relevance of the intervention; 4) Intervention specification via programme theory and a logic model.

**Results:** Narrative programme theory and a simple logic model described ‘*Test to Care*’. It showed the primary intervention function was ‘*incentivisation’,* addressing agency backfill and sick pay. Secondary intervention functions included: ‘*education*’ and ‘*persuasion*’ to staff through communications and social marketing (i.e., posters and emails with embedded short videos); and ‘*environmental/social restructuring*’ and ‘*enablement*’ which invited managers to initiate and support implementation (i.e., the location of testing, management support and poster locations).

**Conclusions:** Our integrative approach to intervention development produced an evidence-based, theoretically-informed intervention tailored to its specific implementation context. The diversity of included inputs were essential to overcome the relative weaknesses and strengths of each input source (e.g., the historical timeframe of published studies in the review, and the sampling biases associated with focus group participation).

**What is already known on this subject?:**

- During the global COVID-19 pandemic, compliance with SARS-CoV-2 testing and other non-pharmaceutical interventions amongst care home staff was enforced through intervention functions such as ‘coercion’ and ‘restriction’ (e.g., mandatory routine testing). However, little is known about how to maintain testing when these intervention functions are removed.
- Intervention development guidance suggests optimal processes should combine the published literature, stakeholder engagement, and programme theory. However, few examples illustrate this process in detail.

**What does this study add?:**

- We present a worked example of using diverse inputs to systematically develop an intervention within a compressed time frame: a typical behaviour change wheel analysis of findings (i.e., barriers and facilitators) from international published studies, novel iterative stakeholder input to add/remove and operationalise intervention content, and sense-checking the emerging intervention’s relevance within the context in which the intervention will be used.
- Uniquely, the study also shows how narrative programme theory and logic models can be used to illustrate an intervention and its purported functions: numerous intervention functions are required (i.e., ‘incentivisation’, ‘education’, ‘persuasion’, ‘environmental/social restructuring’ and ‘enablement’), as are varied behaviour change techniques addressing a range of important mechanisms of action (i.e., ‘knowledge’, ‘beliefs about consequences’, ‘professional role and identity’, ‘social influence’, ‘environmental context and resource’, ‘behavioural regulation’ and ‘memory attention and decision-making’).

## Introduction

The disproportionate mortality rates among residents and staff in United Kingdom (UK) care homes during the COVID-19 pandemic highlighted the extreme vulnerability of the populations residing and working there (e.g., Graham et al., 2020; Gray et al., 2022; Morciano et al., 2021). Beyond COVID-19, interventions are needed to control a wide range of ongoing infections, such as catheter associated urinary tract infections, antimicrobial resistance, influenza, and norovirus (e.g., Gould et al., 2017; Greig and Lee, 2012; Nguyen et al., 2019). As such, care homes represent an important setting where health psychology and behavioural and implementation science are needed to develop, deliver, and optimise interventions that address infectious diseases, as well as a broad range of other health and well-being concerns (e.g., Cooper et al., 2017). To begin the process of addressing this need, here we report on the process of developing an intervention (‘*Test to Care*’) to maintain biweekly asymptomatic SARS-CoV-2 testing amongst English care home staff using lateral flow devices (LFDs) in 2022. The study we describe here was part of a large clinical trial ‘VIVALDI-CT’ (Adams et al., 2023).

### The context

Within the UK, national COVID-19 testing policy was rapidly changing during 2022 (UK Gov, 2022), and it was clear that the evaluation of practicable and scalable interventions was needed to inform future national COVID testing policy (e.g., for the winter of 2023/24). Therefore, between August 2022 and August 2023, VIVALDI-CT – a programme of research with a clinical trial – was funded to address this need. Accordingly, the study was conducted at pace in a highly dynamic environment shaped by changing epidemiology and responsive COVID-19 policy (e.g., UK Gov, 2023).

### Approaches to intervention development

Multiple intersecting and overlapping approaches are available to support the systematic development of health and social care interventions. Each framework can offer a unique contribution to intervention development but they are rarely used in combination to develop optimal interventions. Here, we drew on the Medical Research Council (MRC) complex intervention framework (Skivington et al., 2021); the INDEX study and associated guidance (O’Cathain et al., 2019); programme theory (Pawson and Tilley, 1997); and the behaviour change wheel (BCW) (Michie et al., 2014). Below, we explain how these approaches complement one another (see Figure 1 for a visualisation of their relationships) and each deliver a contribution to the intervention development process.

**Figure 1:**
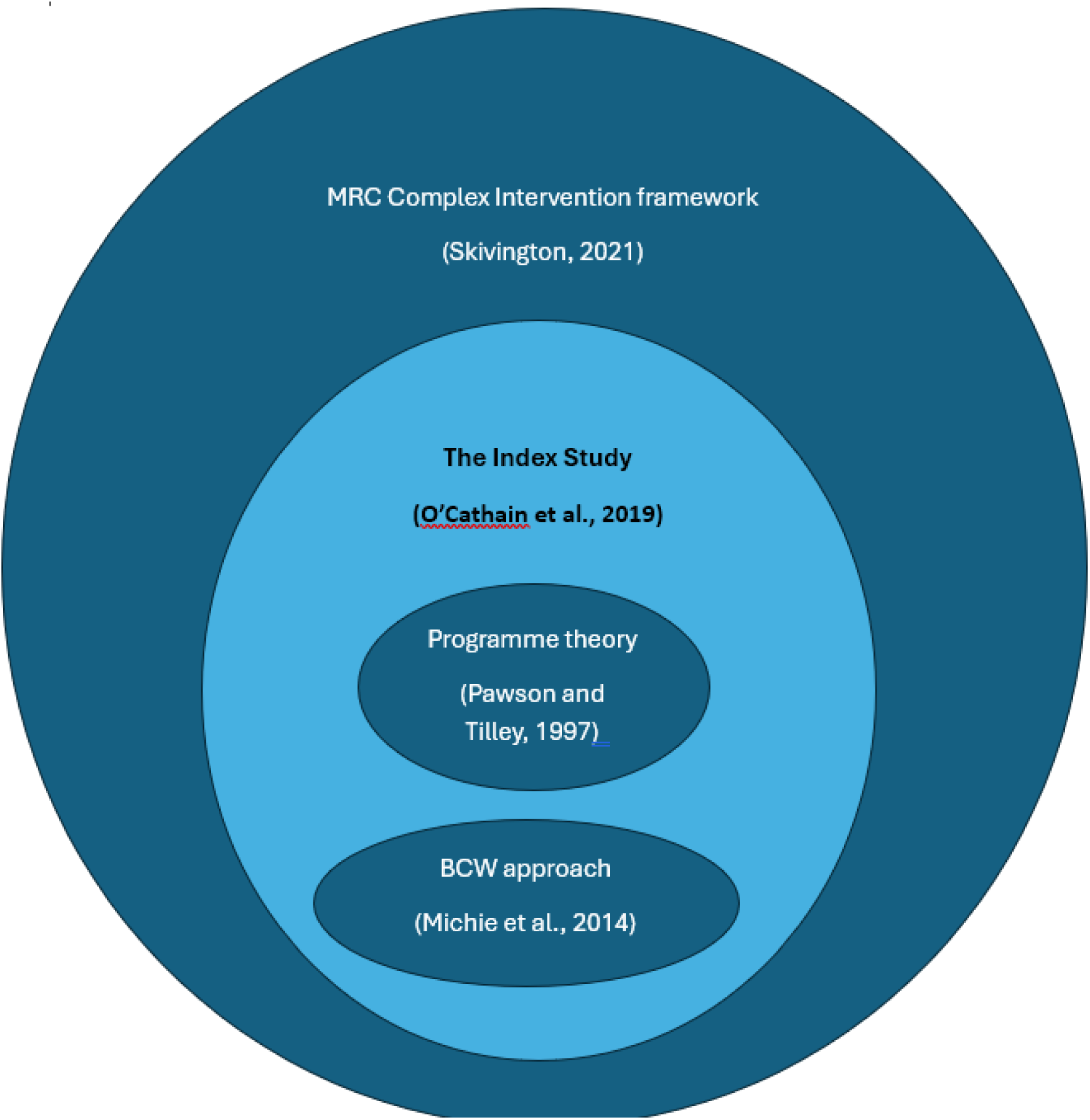
Diverse yet complementary approaches to intervention development.

The MRC Complex Intervention Framework (Skivington et al., 2021) is expansive and encompasses all other approaches but is pitched at a high level. It highlights the cyclical work required to use the best evidence to develop, test, implement, and optimise complex interventions over time and ensure their sustainability and ongoing relevance. In this updated guidance Skivington and colleagues (2021), brought to the fore three novel elements that extend the framework’s previous iterations (e.g., Craig et al., 2007): i) a major focus on understanding how context shapes the way an intervention operates; ii) working continuously with the stakeholders who will be affected by an intervention; and, iii) using programme theory iteratively to assist with initially specifying and then continuously improving the intervention.

Embedded within the MRC framework, further guidance focuses more specifically on processes of intervention development itself:-the INDEX study (‘IdentifyiNg and assessing different approaches to DEveloping compleX interventions’) (O’Cathain et al., 2019). The INDEX study highlights the importance of a) planning the intervention development process; b) reviewing published literature; c) drawing on existing theories; d) undertaking primary data collection; e) paying attention to the future implementation of the intervention in the real world from the very outset of the intervention development process; and f) the principle of designing and refining the intervention continuously.

Similarly cutting across these two approaches ‘Programme theory’ is strongly featured in both the MRC Complex Interventions Framework (Skivington et al. 2021) and the INDEX study (O’Cathain et al., 2019). It is a valuable approach that enables us to design and refine an intervention continuously. It describes the ways we can theorise an intervention working. Drawing on ideas from ‘realist evaluation’ (Pawson and Tilley, 1997), programme theory describes more than just the actual content of the intervention (i.e., what is delivered). It also describes the context in which the intervention is thought to work, the precise ways the intervention works (often referred to as its ‘mechanisms’), and the outcomes the intervention has been designed to change - in trial research, this relates to important measures like the ‘primary and secondary outcomes’.

Programme theory is synonymous with ‘theory of change’ (i.e., de Silva et al., 2014) and both approaches usually draw on visualisations such as ‘logic models’ to communicate an overview of an intervention and its function. Logic models are visual aids used to illustrate, at a high level, how an intervention is thought to work and under which circumstances. There is consensus that programme theory and its associated logic models are best used flexibly during intervention-oriented research. For example, within cycles of intervention development and evaluation, the programme theory will be adapted and updated as new insights into how an intervention works within a given context are more fully understood, or as the intervention and its implementation are optimised based on lessons learned from detailed process and outcome evaluation. Within health psychology however, there are few worked examples of the development of programme theory and associated logic models.

Complementing these relatively broad frameworks of intervention development, more explicitly psychological approaches, such as the BCW (Michie et al., 2014), offer considerable detail for producing interventions to change specific behaviours, building upon decades of previous research and theorisation. The BCW does not provide a wholistic approach to ‘programme theory’ (e.g., context is de-emphasised). However, it represents a highly systematic approach to developing and specifying interventions: describing intervention or implementation *mechanisms* (i.e., what drives behaviour: the Capability, Opportunity, and Motivation (COM-B) approach and/or the Theoretical Domains Framework (TDF); Michie et al., 2011; Carey et al., 2019) and identifying intervention *content* (i.e., what drives behaviour change: intervention functions and behaviour change techniques (BCTs); Michie et al., 2013, 2014).

### The current study

Given the high mortality rates in UK care homes over the COVID-19 pandemic and the lack of research into interventions for infectious disease in this setting, this study aimed to rapidly co-produce a low intensity intervention to maintain biweekly asymptomatic SARS-CoV-2 testing amongst English care home staff. Given this pragmatic goal, we drew on multiple frameworks (See Figure 1), integrated diverse inputs (e.g., See Figure 2), and developed the intervention within a compressed time frame. In this paper, we report the way we developed the intervention, ‘*Test to Care’*, within the English care home sector, as part of the larger clinical trial *‘VIVALDI-CT’* (see Adams et al., 2023). We sought to answer five research questions (RQs):

RQ1) What are key barriers and facilitators to staff testing for SARS-CoV-2 identified from the published literature?
RQ2) What potential intervention content for staff conducting biweekly asymptomatic SARS-CoV-2 is suggested by a BCW analysis of the barriers and facilitators from published literature?
RQ3) How do stakeholders co-produce intervention content for the intervention from the results of the BCW analysis?
RQ4) What are the main barriers and facilitators to staff maintaining biweekly asymptomatic SARS-CoV-2 tests in English care homes just prior to the intervention being delivered, identified from exploratory qualitative interviews?
RQ5) How can the be specified using logic models to depict the underlying programme theory?

**Figure 2.**
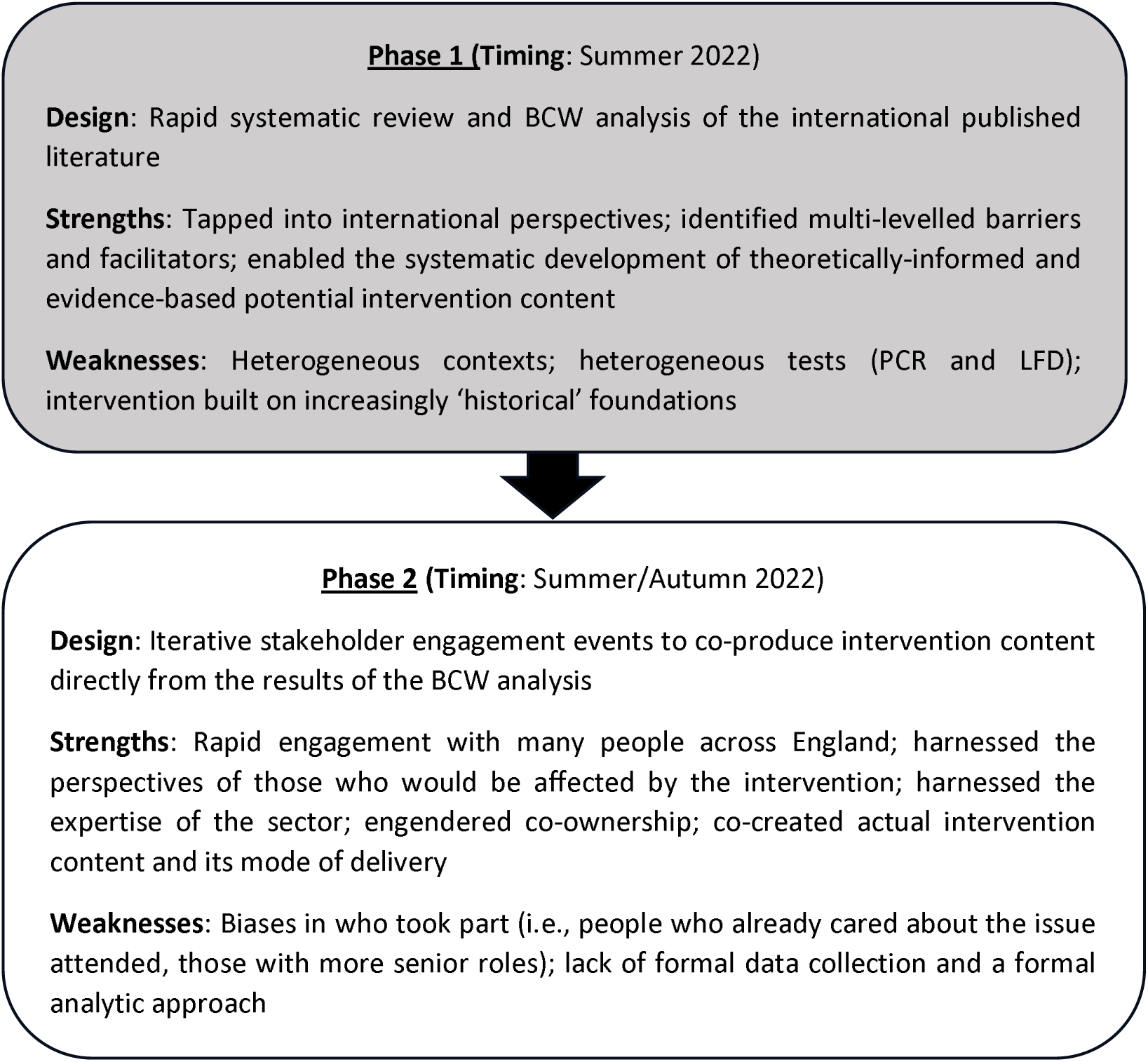
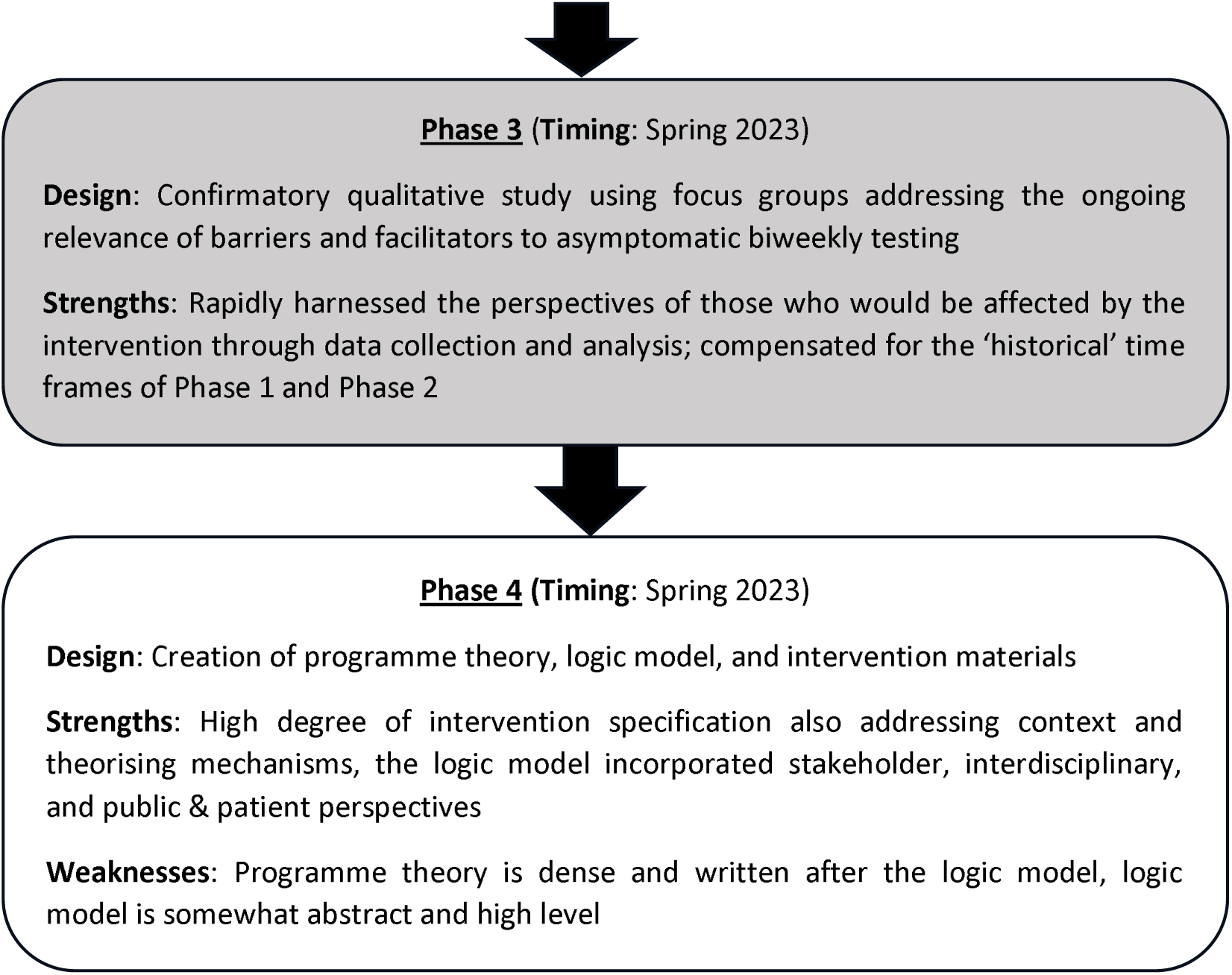
Overall sequential design of the intervention development process.

## Methods

### Overall design

A sequential four-phased, mixed-method design was used to iteratively develop the intervention (see Figure 2). *Phase one* centred on a BCW analysis (Michie et al., 2011, 2014) of the findings of a rapid systematic review of published literature ((for more detail on the systematic review see the protocol-Krutikov et al, 2022) (RQs 1 and 2). *Phase two* consisted of iterative stakeholder engagement events to co-produce the intervention content and delivery plans (RQ3). *Phase three* involved a rapid assessment of the ongoing relevance of barriers and facilitators to asymptomatic SARS-CoV-2 testing that shaped the emerging intervention amongst those who would be directly targeted by the intervention (RQ4). Finally, in *Phase four,* the diverse sources of input were integrated to specify the intervention’s programme theory, visualise it within a logic model, and create intervention resources (RQ5).

#### Phase 1) Rapid systematic review and BCW analysis of the international literature reporting the barriers and facilitators to staff testing for SARS-CoV-2 to generate initial intervention content

##### Rationale

Synthesising published literature to capitalise on what is already known is vital to intervention development (Skivington et al., 2021). The BCW approach enables the systematic generation of potential intervention content from literature concerning barriers and facilitators when existing interventions are unavailable (Michie et al., 2014).

##### Timing

Summer 2022

##### Databases

Ovid MEDLINE, medRxiv, bioRxiv, hand-search references in included review. Search dates were between 2020 and July 2022. Search restrictions were for papers in English language only. The main search terms were: “COVID-19” “care home” or “long term care facility” or “residential home” or “nursing home” and “test” or “screen” or “PCR” or “lateral flow” or “point of care”. As a rapid review intended to develop an intervention in real time, searches were not re-run and quality appraisal of studies was not conducted.

##### Study selection

Identified studies were imported into Mendeley reference management software. Studies were de-duplicated and underwent a three-stage selection process involving title review, abstract view, and full-text review. Study selection and data extraction were performed by four members of the review team (NA, HNL, BA, and MK). Team members worked individually and then cross-checked data extraction. Discrepancies were negotiated.

##### Inclusion criteria

Original research, including observational, interventional, qualitative, and mixed-methods studies, published audits or quality improvement projects, reviews of existing literature, service evaluations, consensus statements, commentaries, and editorials. Studies had to relate to barriers and facilitators to asymptomatic nasopharyngeal screening of SARS-CoV-2 infection in staff working in care homes from UK-comparable settings (i.e., USA, Europe, Canada, Australia, Israel, South Korea, China) between 2020 and July 2022. All types of tests were included.

##### Data extraction

Author (year); country of study; date(s) of study; aims of study; overall design; type of test examined; number of care homes; types of care home; number of participants; and study outcomes.

##### Extraction and analysis of barriers and facilitators

A team of three people (NA, HNL, MK) extracted and standardised data relating to barriers and facilitators using the following construction: “[actor] finds it [easy/hard] to implement asymptomatic testing in [staff] because [reason at a high simple level]”. Subsequently, these standardised barrier and facilitator statements were grouped using the constant comparative method (Grove, 1988) and then categorised using an adaptation of Brofenbrenner’s (1979) ecological systems theory and the socio-ecological model (SEM). Synthesised barriers and facilitators were coded as either relating to the ‘macrosocial’ (care home *sector-level*), ‘mesosocial’ (*care home level*), or ‘microsocial’ (*individual care home staff level)*.

##### BCW analyses

Synthesised barriers and facilitators were further analysed to generate potential intervention content using a series of tools associated with the BCW approach (Michie et al., 2011). These included the TDF to theorise the mechanisms underpinning testing (Cane et al., 2012), corresponding ‘intervention functions’ of the BCW (Michie et al., 2011), and then to specific BCTs using the BCTTv1, Michie et al., 2013). Barrier or facilitator statements clearly relating to uniquely historical factors were excluded from further analysis at this point (e.g., such as shortages of test kits early in the COVID pandemic). These BCW analyses elicited a range of highly specific suggestions for future interventions that could promote the maintenance of COVID-19 testing amongst care home staff. RL led this analysis, and it was checked by PF. Disagreements were resolved through discussion.

#### Phase 2) Iterative stakeholder engagement to co-produce intervention content from the BCW analysis for UK care home staff to maintain biweekly asymptomatic testing for SARS-CoV-2

##### Rationale

The involvement of stakeholders, including those directly affected by an intervention, is good practice for intervention development (e.g., O’Cathain et al., 2019, Preyer et al., 2022, and Skivington et al., 2021). This study took place in fast changing policy context associated with COVID-19. As such Phase 1 had generated evidence-based and theoretically-informed suggestions for potential intervention content, yet this stemmed from international – and increasingly ‘historical’ – contexts. To compensate, it was vital to translate and appraise the suggested intervention content for a contemporary UK context, in addition to working directly with UK stakeholders to learn from them about their ideas for final intervention materials and engender a sense of ownership and ensure cultural relevance.

##### Timing

Summer/Autumn 2022

##### Procedure

A series of eight staggered online engagement events, across the UK care home sector, iteratively operationalised Phase 1 intervention content. Events were run by PF, NA, MK, BA and HN. Groups of stakeholders were presented with the end-results of Phase 1 (e.g., incentives such as payment for COVID-related sick leave, and our ideas around communications/marketing). Stakeholders then shared their perspectives on the ongoing relevance and viability of these ideas, and talked through potential ways that the intervention might be operationalised in practice. We also invited stakeholders to suggest and discuss new intervention content unrelated to the results of Phase 1. Towards the final few events, building on our ongoing learning, we had more detailed discussions about potential intervention names, potential main messages to both managers and staff, the span of potential materials and resources, and ideas about their precise mode of delivery. Throughout, we explicitly sought to tap into stakeholder expertise, elicit their ideas for operationalising intervention content and delivery, and encouraged frank exchange of views (e.g., rejecting ideas from Phase 1 and exploring content suggested in earlier stakeholder events). These events were not recorded or transcribed but notes were taken to track the development and specification of intervention ideas.

##### Participants

Our opportunistic engagement events included a wide variety of stakeholders (e.g., N=∼70) across eight separate events. This events consisted mostly of online meetings with front-line staff and a few care home managers (n=∼40); however, a single meeting with the National Care Forum also took place (an umbrella UK organisation with broad representation from across the care sector) (n=∼20); as well as presentations and discussions with policy makers from across the UK (e.g., representatives from the UK Health Security Agency - UKHSA) and interdisciplinary academics (n=∼10).

#### Phase 3) Rapid confirmatory thematic analysis of barriers and facilitators to UK care home staff maintaining biweekly asymptomatic testing for SARS-CoV-2

##### Rationale

As phase 1 results were based on increasingly ‘historic’ findings from published literature and phase 2 was finalised in the winter of 2022, it was important to confirm its ongoing relevance in the contexts (temporal and cultural) into which the intervention would be implemented. Our rapid study, led by RL, explored the barriers and facilitators to staff testing in English care homes in spring 2023 to precede the roll out of intervention. Analysis focused on confirming the relevance of the meso and micro level barriers and facilitators identified within Phase 1 but was also open to identifying new emerging barriers and facilitators. These analyses were central to considering any further additions, or deletions, to intervention content.

##### Timing

Spring 2023

##### Participants

A diverse convenience sample (N=15) of English staff working in heterogeneous care homes was recruited into four focus groups. Participants were remunerated for their time and expertise. Details of the sample are provided in supplementary file C.

##### Procedure

A brief topic guide encouraged participants to talk about their experiences of testing across the COVID pandemic and the barriers and facilitators to asymptomatic testing. Focus groups were conducted primarily by RL with occasional joint facilitation by PF.

##### Analysis

Transcripts data was deductively thematically categorised by RL and PF to detail barriers and facilitators.

#### Phase 4) Programme theory, logic model, and intervention materials for UK care home staff maintaining biweekly asymptomatic testing for SARS-CoV-2

##### Rationale

To comply with guidance about best practice within intervention development (O’Cathain et al., 2019) and future evaluation, it was important to describe the intervention and how it was thought to work then develop concrete ways of delivering the intervention to be implemented within the trial.

##### Timing

Spring 2023

##### Analysis

RL and PF drafted the logic model, incorporating details of what was known about the context, the specific problem the intervention was designed to address, the intervention content that would be delivered (i.e., Phase 2), the intervention mechanism (e.g., drawing on the results of the BCW analyses in Phase 1) and the intervention outcomes. The logic model was sense-checked and refined with both interdisciplinary colleagues (n=8) and PPIE representatives (N=8) to ensure that there was consensus over it reflecting the overall intervention and the way it was imagined to work, and that it also made sense as a communication device to be used with diverse audiences. Subsequently, PF led the development of a more expansive narrative programme theory to provide a detailed account of how the intervention was thought to work. This was audited and checked by RL and a range of interdisciplinary peers many of whom were not involved with the project (n=∼15).

The study received ethical approval from the London - Bromley Research Ethics Committee (22/LO/0846) and Health Research Authority Confidentiality Advisory Group approval (22/CAG/0165)

## Results

### Phase 1) Rapid systematic review and BCW analysis of barriers and facilitators to care home staff maintaining testing for SARS-CoV-2 from international literature to generate initial intervention content

Details of the 14 methodologically diverse studies identified in the rapid systematic review are reported elsewhere (in supplementary files A). Five studies were from the UK, five from the USA, four from the rest of the world. They were of varying designs and involved either PCR or LFD tests for SARS-CoV-2. Here, we concentrate on reporting the multileveled barriers and facilitators to testing within the literature and the end results of our BCW analysis (see Table 1 which includes details of their frequency). Details of the underpinning BCW analysis are available elsewhere (in supplementary files Table B).

**Table 1.** An overview of standardised and synthesised barriers and facilitators to COVID testing in care home and the results of the BCW analyses.

| **BARRIER** | **#\*\*** | **FACILITATOR** | **#\*\*** | **Suggested Intervention content resulting from the BCW analysis\*** |
| --- | --- | --- | --- | --- |
| **Macrosocial (care home sector-level)** |  |  |  |  |
| The care home sector found it hard to implement testing because of the financial costs involved | 1 | The care home sector found it easy to implement testing when providers received financial reimbursement for staff absence | 1 | Consider means of manipulating payment as intervention content [14, 25] |
| *The care home sector found it hard to implement testing when guidance was changing, conflicting, hard to interpret or unavailable* | 6 | *The care home sector found it easy to implement testing when guidance was clear, coherent, and consistent* | 2 | *Historical findings relating to the changing guidance in many countries during the COVID-19 pandemic - not amenable to change in future interventions* |
| *The care home sector found it hard to implement testing because care homes can differ greatly from each other and national guidance didn't always apply to individual settings* | 2 | *The care home sector found it easy to implement testing when there were protocols already in place that could be activated when need be* | 2 | *Historical findings relating to the COVID-19 pandemic and the care home sectors struggles to engage and interpret changing guidance - not amenable to change in future interventions*<br>*Consider the heterogeneity of diverse care homes and include clear planning and protocols for testing [23]* |
|  |  | *The care home sector found it easy to implement testing when there were pre-established partnerships between homes and external organisations (e.g. clinical, laboratory, public health)* | 3 | *Historical findings relating to the changing guidance in many countries during the COVID-19 pandemic - not amenable to change in future interventions*<br>*Consider capitalising upon these relationships when designing future intervention content [21]* |
| *The care home sector found it hard to implement testing because tests were not always readily available* | 1 | *The care home sector found it easy to implement testing when testing kits are readily available and free of charge* | 1 | *Historical findings relating to the early days of the COVID-19 pandemic when tests were being developed and were not consistently available - not amenable to change in future interventions* |
|  |  | Care home sector finds it easy to implement asymptomatic testing because it is framed as being part of a wider health promotion effort | 1 | Historical findings relating to the early days of the COVID-19 pandemic - not amenable to change in future interventions |
|  |  | Care home sector finds it easy to implement asymptomatic testing when the sector was held accountable for new infections | 1 | Historical findings relating to the early days of the COVID-19 pandemic - not amenable to change in future interventions |
| **Mesosocial (care home level)** |  |  |  |  |
| Care homes found it hard to implement testing due to concerns about staff shortages in relation to positive results | 6 | Care homes found it easy to implement testing when additional staff could be deployed to backfill for COVID related staff absences | 2 | Consider using resource to provide back fill for staff with positive results (12.1 and 12.2) delivering social support (3.1 and 3.2) and work with care homes to resolve staff shortages locally (1.2) [32, 57] |
| Care homes found it hard to implement testing due to the additional administrative workload it required | 7 | Care homes found it easy to implement testing when there were plans in place for this eventuality | 3 | When planning an intervention consider changing the timing and location of testing (12.1 and 12.2) outwith the workplace and also encourage local solutions (1.2) to minimise administrative strain wherever possible (12.1) [34,59] |
|  |  | Care homes found it easy to implement testing when plans were communicated well to staff | 3 | Ensure that plans are well communicated to staff - using a credible source to make clear their role and responsibilities (5.1, 5.3, 9.1, 2.7, 12.2) [61,59] |
| Care homes found it hard to implement testing as a result of poor and/or inconsistent training | 2 |  |  | Provide on-going and accessible training for staff that shows how to test (6.1,4.1) and encourages practice (8.1) [36, 94, 96] |
|  |  | Care homes found it easy to implement testing when there was ongoing consultation with those involved in the testing procedure | 2 | Ensure ongoing consultation with those involved in testing, in order to gauge opinions/outlooks (5.3, 5.6, 6.3, 2.2) and consider workarounds/improvements (1.2) [63] |
| Care homes found it hard to implement testing due to the intense | 1 | Care homes found it easy to implement testing because of the | 1 | Ensure communications and marketing reinforce and stress the positive health and social consequences of |
| frequency and regularity with which it was required |  | belief that testing was/is important and valuable |  | testing (5.1, 5.3) [53, 55] |
|  |  | Care homes found it easy to implement testing when staff had pre-existing relationships with the residents | 1 | Ensure communications and marketing reinforce and stress the positive health and social consequences of testing (5.1, 5.3) [53, 55] |
| Care homes found it hard to implement testing due to concerns about the reliability of testing kits and their subsequent results | 1 | Care homes found it easy to implement testing because of the belief that the approach to testing was effective and practical. | 1 | Ensuring communication and marketing includes a credible source (9.1) that endorses testing whilst educating and persuading staff about the value of testing (5.1) and the positive impact it can have above and beyond other IPC measures (5.3) [42,67] |
| Care homes find it hard to implement asymptomatic testing because of the potential reputational damage done to homes which are seen to have high numbers of positive cases | 1 |  |  | Ensure communication and marketing persuade care home staff to test through a credible source (9.1) which focuses on the goals of keeping people safe and the care home open by stressing past success (15.3, 2.7) and highlighting the consequences of testing and not testing (5.1, 5.3). [46] |
| *Care homes found it hard to implement testing when there were issues with the home having access to adequate numbers of testing kits* | 1 |  |  | *Historical findings relating to the early days of the COVID-19 pandemic when tests were being developed and were not consistently available - not amenable to change in future interventions* |
| *Care homes found it hard to implement testing due to the impact of lack of space on standardised pathways for testing* | 3 | *Care homes find it easy to implement asymptomatic testing when the protocol for staff testing and resident testing is kept separate and distinct* | 1 | *Historical stemming from the demands of social distancing whilst testing and testing both residents and staff at the same time - not amenable to change in future interventions* |
| *Care homes found it hard to implement testing because of delays in test results* | 1 |  |  | *Historical stemming from the use of PCR tests before LFD were available for rapid results - not amenable to change in future interventions* |
| *Large care homes found it hard to implement testing due to their size* | 1 |  |  | *Not amenable to change in future interventions* |

| **Microsocial (Individual care home staff level)** |  |  |  |  |
| --- | --- | --- | --- | --- |
| Care home staff find it hard to implement asymptomatic testing because of the financial implications of testing positive and being off sick | 1 | Care home staff found it easy to implement testing because they viewed it as helping to prevent a resurgence of COVID-19 and the risk of going back into lockdown | 1 | Ensure communication and marketing persuade and incentivise care home staff through fiscal change and marketing about the removal of financial barriers to testing (5.3) [70]<br>Ensure communication and marketing persuade care home staff to test through focusing on the goals of keeping people safe and the care home open by stressing past success (15.3, 2.7) and highlighting the consequences of testing and not testing (5.1, 5.3). It may also be useful to consider drawing on identities associated with changed behaviour (13.5) [103] |
|  |  | Care home staff found it easy to implement testing because they took pride in caring for residents, and keeping them safe, to the best of their ability | 1 | Ensure communication and marketing persuade care home staff to test through speaking directly to their professional identity and professional pride - consider the use of self as a role model (13.1) or content that shows the varied content of testing or not testing (5.1, 5.3, 5.6, 5.2) [113] |
| Care home staff found it hard to implement testing because of concerns about staff capacity due to positive results | 1 |  |  | Consider using resource to provide back fill for staff with positive results (12.1 and 12.2) delivering social support (3.1 and 3.2) and work with care homes to resolve staff shortages locally (1.2) [32, 57, 74] |
| Care home staff found it hard to implement testing because the process added considerably to their workload | 6 | Care home staff found it easy to implement testing because they viewed its benefits as outweighing its costs | 1 | When planning an intervention consider changing the timing and location of testing (12.1 and 12.2) outwith the workplace and also encourage local solutions (1.2) to minimise workload strain wherever possible (12.1) [84] |
| Care home staff found it hard to implement testing because they felt overworked and under-appreciated within the care sector | 2 |  |  | Ensure communication and marketing persuade care home staff to test through speaking directly to their professional identity and professional pride - consider the use of self as a role model (13.1) or content that shows the varied content of testing or not testing (5.1, 5.3, 5.6, 5.2) [90] |
| Care home staff found it hard to implement testing because they didn't believe it was a valuable addition to their usual practice | 1 | Care home staff found it easy to implement testing because they viewed it as a useful and valuable addition to their usual practice | 3 | Ensuring communication and marketing includes a credible source (9.1) that endorses testing whilst educating and persuading staff about the value of testing (5.1) and the positive impact it can have above and beyond other IPC measures (5.3) [76] Ensure communications and marketing stress the positive health and social consequences of testing (5.1, 5.3) [105] |
|  |  | Care home staff find it easy to implement asymptomatic testing because it enables external support from other HCPs - i.e. GPs, health visitors - which lockdown prevented | 1 | Ensure communications and marketing reinforce the positive outcomes associated with relaxation of COVID-19 precautions (1.4, 12.1) - especially focusing on the facilitation of external support from other HCPs (3.2, 12.2) [107] |
| Care home staff found it hard to implement testing because they had concerns about the efficacy of the testing kits | 3 | Care home staff found it easy to implement testing because they believed that the testing kits were effective in provide accurate results | 1 | Ensuring communication and marketing includes a credible source (9.1) that endorses testing whilst educating and persuading staff about the value of testing (5.1) and the positive impact it can have above and beyond other IPC measures (5.3) [80] |
| Care home staff found it hard to implement testing because of the substantial administrative strain it added to their workload | 2 | Care home staff found it easy to implement testing when their managers endorsed testing, and supported & enabled them to be able to do so effectively | 3 | When planning an intervention consider changing the timing and location of testing (12.1 and 12.2) outwith the workplace and also encourage local solutions (1.2, 12.1) [82, 98, 101] |
| Care home staff found it hard to implement testing because testing on-site required them to travel to work outside of their scheduled hours and on days off | 4 |  |  | When planning an intervention consider how care home staff management structures work and modify them (12.1 and 12.2) to ensure that managers endorse and support testing (9.1, 3.2) with regular prompts to |
| Care home staff found it hard to implement testing because of the enormity of planning required to carry it out | 1 |  |  | test (7.1). It may be useful to consider the influence of managers evoking a professional responsibility and duty of care (6.2, 6.3) [101] |
| Care home staff found it hard to implement testing because of poor communication from management about guidelines, responsibilities, and expectations | 1 |  |  | Ensure communications and marketing reinforce the positive outcomes associated with relaxation of COVID-19 precautions (1.4, 12.1) [107] |
| Care home staff found it hard to implement testing because they lacked skills and were not adequately trained | 6 |  |  | Provide on-going and accessible training for staff that shows how to test (6.1,4.1) and encourages practice (8.1) [94] |
| Care home staff find it hard to implement asymptomatic testing because of concerns about the cost of testing kits | 1 |  |  | Ensure communication and marketing persuade and incentivise care home staff through fiscal change and associated marketing about the removal of financial barriers to testing (5.3) [72] |
| *Care home staff found it hard to implement testing because they didn't have access to testing kits* | 2 |  |  | *Historical findings relating to the early days of the COVID-19 pandemic when tests were being developed and were not consistently available - not amenable to change in future interventions* |
| *Care home staff find it hard to implement asymptomatic testing because of logistical difficulties in sending and/or receiving results* | 4 |  |  | *Historical stemming from the use of PCR tests before LFD were available for rapid results - not amenable to change in future interventions* |

Note. \*See Table B in the supplementary files for full BCW analysis. \*\*# = Number of times barrier/facilitator appeared in papers. Numbers in brackets e.g., '(7.1)' relate to the specific behaviour change techniques (BCTs) identified using the BCTTv1 within the BCW analysis shown in Table B in the supplementary files. Numbers in square brackets relate to the row number of Table B in the supplementary files. Italics denote historical findings and barriers/facilitators that are not amenable to change in future interventions.

Using the SEM, at the *macrosocial* level, 21 barriers and facilitators were identified which related to testing *across the care home sector* (e.g., within the whole care home system of multiple and diverse organisations). From these, 15 were not amenable to change within future interventions because they were historic and no longer relevant and related to the very early period of the COVID-19 pandemic (see Table A). Such historic findings focused on issues like labile policy, guidance, and the availability of COVID tests. These were not taken forward for further BCW analysis. However, the role of financial reimbursements for COVID-related absences in driving testing was an important finding. The BCW analysis (Table B) suggested manipulating financial reimbursement for positive results may be an important intervention function (‘incentivisation’) in supporting testing maintenance.

At the *mesosocial* level, 38 reported barriers and facilitators to testing *within care homes* were identified (e.g., within care homes as distinct individual organisations). Seven were not amenable to change as they were historical. These were not taken forward for further BCW analysis. Included barriers comprised issues of administrative workload associated with testing (particularly when it was on-site), training about testing, the frequency and regularity of testing, and collective concerns about the reliability of tests. Facilitators included the role of planning, good communication within homes, collective beliefs in the value of tests (i.e., that testing can reduce infections), longstanding relationships between staff and residents, and collective beliefs in the effective nature of tests. The BCW analysis suggested that using resources to provide back fill for staff with positive results could be important, as could changing the location of testing from within care homes to staff homes; that communicating plans to staff would be central; and that training should be accessible and ongoing - encouraging practice and showing how to test. The BCW analysis also highlighted the role of communications and social marketing from a credible source to educate and persuade care homes about the value and positive consequences of maintaining testing.

At the *microsocial* level, 43 barriers and facilitators to testing *for individual care home staff* (e.g., within individual care home workers) were identified, eight of which were not amenable to change. These were not taken forward for further BCW analysis Included barriers and facilitators often resonated with the macro- and mesosocial issues outlined above. Barriers included the financial implications of testing and concerns about staff capacity if test results were positive leading to staff sickness absence, concerns about additional test-related workload, feelings about their sector being under-appreciated, concerns about the added value of testing and its efficacy, travel costs associated with testing in the workplace, the need for planning, poor communication from management about responsibilities, and a lack of adequate training. Facilitators included beliefs in the value of testing to prevent a resurgence in COVID; pride in keeping residents safe; beliefs that the benefits outweighed the costs and in its added value above and beyond other infection prevention and control measures; beliefs in the efficacy of testing; and support and endorsement from management. The associated BCW analysis suggested future intervention content should ‘incentivise’ and ‘persuade’ staff to test through the removal of financial barriers to testing, harness the goals of keeping care homes open and residents safe, highlight the consequences of testing and not testing, consider the time and location of testing outwith the workplace, speak to staff’s professional identity, incorporate a credible source within communications that emphasise the added value of testing and the test kit efficacy, ensure that local management structures endorse and support testing, consider regular prompts to testing, evoke a professional responsibility and duty of care, ensure communications reinforce the positive outcomes associated with the relaxation of COVID precautions, and ensure ongoing and accessible training.

### Phase 2) Iterative stakeholder engagement events to co-produce intervention content from the results of the BCW analysis

Table 2 provides an overview of the end result of the stakeholder co-production process. The left-hand column shows suggested intervention content from Phase 1. The middle column shows the range of intervention ideas which were rejected (at the macro, meso, and micro levels), including some new ideas suggested within the engagement events themselves. In the right-hand column, we show those ideas which were maintained and further developed across these diverse events.

**Table 2.** An overview of intervention content for UK care home staff maintaining biweekly asymptomatic testing for SARS-CoV-2 as co-produced by stakeholders.

| BCW intervention content* and<br>*New content suggested by stakeholders in italics* | Ideas not to include in the intervention | Ideas to include in the intervention |
| --- | --- | --- |
| **Macrosocial (care home sector-level)** |  |  |
| • Consider means of manipulating payment as intervention content [25]<br>• *Policy changes for intervention homes should incentivise testing through the removal of face masks*<br>• *Policy changes for intervention homes should use confirmatory PCR testing* | • Policy changes for intervention homes should include payment for the testing process in addition to sick pay for positive results<br>• *Policy changes for intervention homes should incentivise testing through the removal of face masks*<br>• *Policy changes for intervention homes should use confirmatory PCR testing* | • Intervention homes should receive sick pay for those testing positive as this could have major policy implications for the UK<br>• Intervention homes should receive payment for agency staff to ensure homes with staff testing positive still function<br>• Policy changes for intervention homes should include the ongoing provision of free LFD test kits to ensure that this is not a barrier to testing |
| **Mesosocial (care home level)** |  |  |
| • When planning an intervention, consider changing the timing and location of testing (12.1 and 12.2) outwith the workplace and also encourage local solutions (1.2) to minimise administrative strain wherever possible (12.1) [34]<br>• Ensure that plans are well communicated to staff - using a credible source to make clear their role and responsibilities (5.1, 5.3, 9.1, | • Care homes should mandate the location of testing<br>• Care homes should implement local testing champions (managers or peers) | • Care homes should implement local solutions to location of testing<br>• Use existing structures to cascade the intervention and its main messages |
| 2.7, 12.2) [61]<br>• Provide on-going and accessible training for staff that shows how to test (6.1,4.1) and encourages practice (8.1) [94/96] | • Care homes should implement training and monitoring for LFD use<br>• Care homes should implement the auditing of LFD techniques | • Care homes should offer access to existing training for correct LFD use if it was required |
| **Microsocial (Individual care home staff level)** |  |  |
| • Ensure communication and marketing persuade care home staff to test through focusing on the goals of keeping people safe and the care home open by stressing past success (15.3, 2.7) and highlighting the consequences of testing and not testing (5.1, 5.3). It may also be useful to consider drawing on identities associated with changed behaviour (13.5) [103]<br>• Ensure communication and marketing persuade care home staff to test through speaking directly to their professional identity and professional pride - consider the use of self as a role model (13.1) or content that shows the varied content of testing or not testing (5.1, 5.3, 5.6, 5.2) [90]<br>• Ensuring communication and | • Naming of intervention: 'Pay for Tests', 'Test for Now', 'Flare to care', 'Keeeeeep testing', 'Extra Testing'<br>• A main focus on professional pride and identity and career recognition<br>**Modality of intervention delivery:** Short videos featuring carers to promote the intervention and be emailed or uploaded to chain websites (brand, tag line, main messages), 'Newsletter' for managers/staff, Ready constructed reminders for managers to send out (scripted main messages with BCTs)<br>**Main messages to staff:** 'Reminders to test twice a week when without symptoms from peers/managers', 'Regular communications between staff will be encouraged', 'Balance humour | Naming of intervention: 'Test to care'<br>**Modality of intervention delivery:** Test kits provided continuously, Poster(s)- positioned near clocking in machine and around the care homes, Email/text content for managers (main messages), Payment/transfer system for £, Short videos featuring expert to promote the intervention -to be emailed<br>**Main messages to staff:** 'Support your peers with their testing and their recording of results', 'Training is always available - if you need a reminder or the kit has changed', 'Test where and when your manager tells you to', 'provide access to LFD training for those |
| marketing includes a credible source (9.1) that endorses testing whilst educating and persuading staff about the value of testing (5.1) and the positive impact it can have above and beyond other IPC measures (5.3) [76] Ensure communications and marketing stress the positive health and social consequences of testing (5.1, 5.3) [105]<br>• When planning an intervention consider how care home staff management structures work and modify them (12.1 and 12.2) to ensure that managers endorse and support testing (9.1, 3.2) with regular prompts to test (7.1). It may be useful to consider the influence of managers evoking a professional responsibility and duty of care (6.2, 6.3) [101] | with health', 'Directly address 'care home exceptionalism', 'Badges with intervention messages', 'Purpose of trial within brand'<br>**Tag lines:** 'Test twice a week for a better future', 'Do extra testing to protect all', 'Testing, trust, teamwork', 'Be the best – do two tests', 'Doing things together - for each other', 'Testing together: second nature', 'Flaring is caring (with a big nose cartoon)', 'Remember how bad it was: flare to care', 'You never “nose” what's up your nose', 'Stop, think, test! Traffic light system', 'Be the best do a test'<br>**Main messages to managers:** Send reminder texts, Spot check on bar codes of LFD results, Make use of mentors for new staff | who may need it'<br>**Tag lines:** "Test with peace of mind knowing you'll get sick pay if your results are positive", "Doing extra testing helps us protect each other- Staff and residents"<br>**Main messages to managers:** Build on your past experience/expertise, Develop a system that guides staff about which days to do their tests (when they are most likely to have been exposed), Develop a local system to check authenticity of results, if you feel you need to, Develop a system that supports managers making payments that works locally, Remind your staff about testing twice a week and recording the results, Support your staff with testing twice a week and recording the results, Regular communications about testing and recording between managers and staff |

*Note.* \*See Table B in the supplementary files for full BCW analysis. Numbers in brackets e.g., (7.1) relate to the specific behaviour change techniques (BCTs) identified using the BCTTv1 within the BCW analysis shown in Table B in the supplementary files. Numbers in square brackets relate to the row number of Table B in the supplementary

Notable rejected intervention ideas included the stakeholder-generated idea of the behavioural substitution of ‘testing’ for ‘mask-use’ (at the time staff mask use was mandatory in UK care homes). This was rejected because of the regulatory complexity of implementing such an idea at scale. Other notable rejected ideas included the use of testing champions – this had been unsuccessfully attempted within the COVID pandemic and was too intensive for a pragmatic scalable intervention, as well as being potentially irritating to staff. Instead, cascading messages within existing management structures was agreed as a useful compromise to reinforce the intervention if and when it was required as testing or COVID policy changed.

Ideas about the provision of ongoing training, monitoring, and auditing of testing were also rejected, given the high level of testing expertise in this sector and the habitual nature of regular testing for most care home staff and their strong desire to protect residents and colleagues. In relation to the communications and marketing, there was a sense that this needed to strike the right balance between being directive but not patronising or abusive. Although the use of humour within the visual materials had been suggested (e.g., ‘Flare to care’, ‘Keeeeeep testing’), it was eventually rejected as high-risk given the trauma some staff and homes had been through during the COVID pandemic. Equally, manipulating ‘professional role and identity’ was rejected given how many staff felt this classic mechanism of action (Atkins et al., 2017) had been overused during the peak of the COVID pandemic and could be seen as ‘emotional blackmail’.

Table 2 also shows the intervention ideas which were kept, developed further, and operationalised. Notable components included the financial mechanisms to cover sick pay for positive test results. In relation to social marketing and communications, the name ‘Test to Care’ was well-liked by many. Over time and across the various events, most stakeholders agreed that a light-touch approach to LFD training and auditing – with reminders about the need to test and upload test results – was the best approach to encourage maintenance of testing; it was suggested that more heavy-handed approaches would be a further reminder to staff of ‘being made to do things’, which had been imposed on both them and the care home sector.

### Phase 3) Rapid confirmatory thematic analysis of barriers and facilitators to UK care home staff maintaining biweekly asymptomatic testing for SARS-CoV-2

Table 3 (above) shows the ongoing relevance of the barriers and facilitators identified within the earlier systematic review. This supported for the planned intervention working as anticipated within the UK care home context in 2023. Findings are presented in order of decreasing frequency with indicative illustrative data extracts. All the main barriers and facilitators within the systematic review were present within the data.

**Table 3.** An overview of barriers and facilitators to UK care home staff maintaining biweekly asymptomatic testing for SARS-CoV-2 identified from thematic analysis.

| BARRIER | ## | FACILITATOR | ## | INDICATIVE QUOTE | Implications for intervention content |
| --- | --- | --- | --- | --- | --- |
| **Mesosocial (Care home level)** |  |  |  |  |  |
| Care homes found it hard to implement testing due to the additional administrative workload it required | 14 | Care home staff found it easy to implement testing when their managers endorsed testing, and supported & enabled them to be able to do so effectively | 10 | 'You can just do it in your own time. You had peace of mind because at 9 o'clock in the morning, you can see that you're COVID-free, and then you can relax.. and then you just get to your shift and carry on. Rather than having to get to work 20 minutes earlier, and people are trying to talk to you, and bells are ringing, but you're still trying to record [your result]'. [FG3, 011] | Confirmed ongoing relevance of these barriers and facilitators and supported the planned intervention content |
| Care home staff found it hard to implement testing because the process added considerably to their workload | 5 | Care homes found it easy to implement asymptomatic testing because they developed structures and processes around testing to improve compliance | 5 | 'We had a designated person that would be in the room all day doing the testing and registering them. I'd be in that room all day doing the tests, and then registering all the staff and residents as well. So it was a several hour job... it was an inconvenience, but a necessary evil I guess' [FG3, ppt002] | Confirmed ongoing relevance of these barriers and facilitators and supported the planned intervention content |
| Care home staff found it hard to implement testing because they were not adequately trained | 5 | Care home staff found it easy to implement testing because they were adequately trained | 2 | 'Obviously training is really important for the testing staff. Even if we're doing it again and again consecutively, if we don't know the proper way of doing it then every time we're doing it wrong. So training, I think, is really essential' [FG3, 004] | Confirmed ongoing relevance of these barriers and facilitators and supported the planned intervention content |
| *Care homes found it hard to implement* | 3 | *Care homes found it easy to implement asymptomatic* | 3 | 'The easiness of the testing is because of the availability of the test kits – if the | New finding relating to the changing policy context with |
| *asymptomatic testing because staff didn't have adequate access to testing kits\** |  | *testing because testing kits were readily available to staff* |  | kit is no longer available, people will not test again. They should make sure every home has kits available' [FG1, 001] | implications to ensure free test kits are always available |
|  |  | *Care homes found it easy to implement asymptomatic testing because for staff it had become routinised and 'second nature'* | 6 | 'I think what we experienced is that it became second nature for us to go through all these testings and now we... the masks have been taken away and the testing has been reduced and it feels like we've lost a leg and an arm. It's like... because it's not part of the routine, something's missing' [FG2, 011] | New findings but they confirmed the ongoing relevance of these barriers and facilitators and supported the planned intervention content |
| **Microsocial (Individual/Care home staff level)** |  |  |  |  |  |
| Care home staff found it hard to implement testing because of the financial implications of testing positive | 15 | Care home staff found it easy to implement testing because of the financial implications of testing positive | 4 | 'If people aren't getting paid, then they aren't going to test, because they just can't afford to be off work' [FG1, ppt005] | Confirmed ongoing relevance of these barriers and facilitators and supported the planned intervention content |
| Care home staff found it hard to implement asymptomatic testing because they had concerns about the efficacy of the testing kits | 12 | Care home staff found it easy to implement asymptomatic testing because they believed that the testing kits were effective in providing accurate results | 7 | 'I went to work... had an LFD, that was negative... started feeling unwell.. did another one and that was positive, and my PCR came back positive. So they're a bit unpredictable' [FG1, 003] | Confirmed ongoing relevance of these barriers and facilitators and supported the planned intervention content |
|  |  | Care home staff found it easy to implement testing because they took pride in caring for residents, and keeping them safe, to the best of their ability | 13 | 'the only thing that helps with testing is knowing that we are helping to keep other people virus-free – that's the only thing that matters for me. [...] The only reason you're doing it is because there's other lives at stake. If it was up to me, I wouldn't test – because we've been doing it for two years, sometimes twice a day – so no, the only thing that keeps you going is the fact you're doing it for other people' [FG2, 011] | Confirmed ongoing relevance of these barriers and facilitators and supported the planned intervention content |
| *Care home staff found it difficult to implement asymptomatic testing because of the physical discomfort involved* | 5 | Care home staff found it easy to implement asymptomatic testing because they viewed its benefits outweighed its costs | 5 | 'What makes it so difficult is having to insert in into your nose.. it's so uncomfortable, so sometimes people will tend not to do it correctly. Most times it can be traumatic to actually go for it due to the associated pains involved' [FG1, 001] | Some new findings but overall confirmed ongoing relevance of these barriers and facilitators and supported the planned intervention content |
|  |  | *Care home staff found it easy to implement asymptomatic testing because testing was simply a prerequisite of their job and they had no choice in the matter* | 7 | 'If our management or the government says 'test', then we just do it. So any name [re branding/messaging] that's put to it, needs to encourage you to do it, but I think where we come from, it's just sort of like we have to do it. We don't have a choice. There is no 'we don't feel like doing this today' – in other environments maybe yes – but we don't have that choice' [FG2, 011] | New findings – highlighting the need for persuasive communications |
| Care home staff found it hard to implement asymptomatic testing because they didn't believe it was a valuable addition to their usual practice | 1 | Care home staff found it easy to implement testing because they viewed it as a useful and valuable addition to their usual practice | 4 | 'Effective communication and public awareness can be good education. Educating people about the importance of testing is really important for them, and also at the same time dispelling any myths and addressing any concerns that people are facing, and providing the materials and convenience to people will encourage testing... and obviously the communication coming from a trusted source like the NHS – if they tell the people to test it will increase the number of people testing' [FG2, 004] | Confirmed ongoing relevance of these barriers and facilitators and supported the planned intervention content |
|  |  | *Care home staff found it easy to implement asymptomatic testing because it was completely routinised* | 5 | 'It's become a normality. So it's when you get any symptoms as well now, any sort of COVID-related symptoms, you just reach for an LFD' [FG3, 002] | Some new findings relating to the changed context- however the main elements of the planned intervention should remain salient |
| *Care home staff found it difficult to implement asymptomatic testing because of the reduced severity of symptoms and changing societal attitudes* | 2 | *Care home staff found it easy to implement asymptomatic testing because the COVID-19 pandemic increased their anxiety about their health generally and increased their vigilance in seeking to identify positive cases* | 2 | 'when stuff started to calm down and you were still having to test 3 times a week, it felt like a bit of a slap in the face when people were saying 'well I can go out and do whatever I want' ... that was sometimes hard for staff, wanting a bit of a normal life after what you've gone through ... no-one else has had to do any of this stuff, but we still have to do it, I think that was one of the things that people struggled with' [FG1, 004] | Some new findings relating to the changed context- however the main elements of the planned intervention should remain salient |

*Note.* \*Text in italics signifies new findings not detailed within the systematic review. \*\*The numbers within the two columns indicate a frequency count of these barriers and facilitators being mentioned

Additional barriers and facilitators were identified. These included that participants acknowledged the importance of access to testing kits to enable testing. They also talked about how testing had become routinised over the course of the COVID pandemic, facilitating the imagined maintenance of biweekly testing. New findings also identified the role of pain and irritation as a barrier to ongoing testing, managerial direction enabling ongoing testing, and new challenges to testing having emerged as the risk of severe outcomes in residents following a positive test diminished substantially after vaccination, reducing the perceived benefits of the intervention. There was also a growing disparity with what staff in care homes were being asked to do and what was happening in other health and social care contexts (e.g., mask use was mandatory in care homes but not within NHS settings).

### Phase 4: Creation of programme theory, logic model, and intervention materials

Figures 3,4 and 5 show the development of the logic model depicting the development of the intervention.

**Figure 3.**
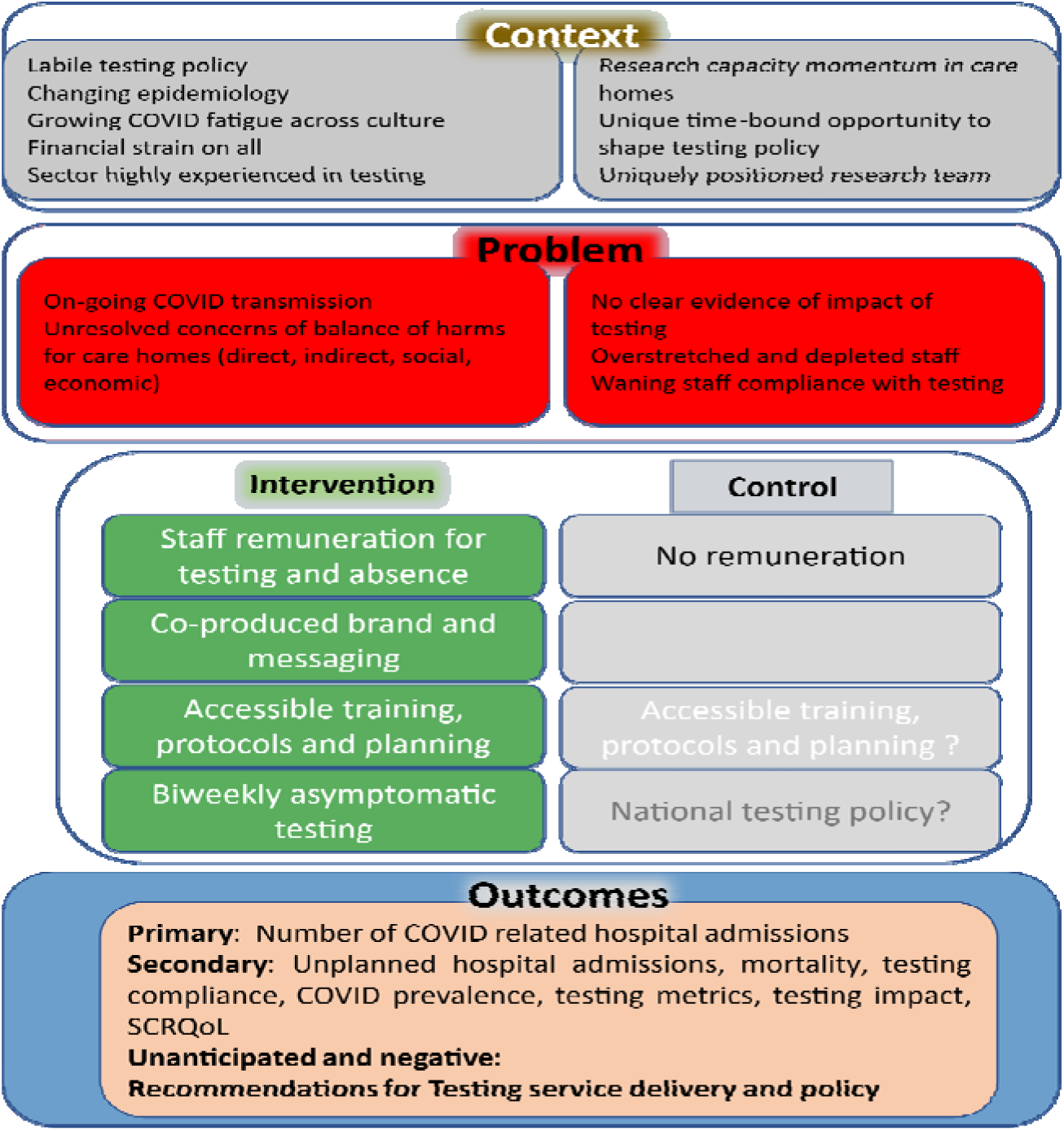
Initial logic models showing basic model of the intervention.

**Figure 4.**
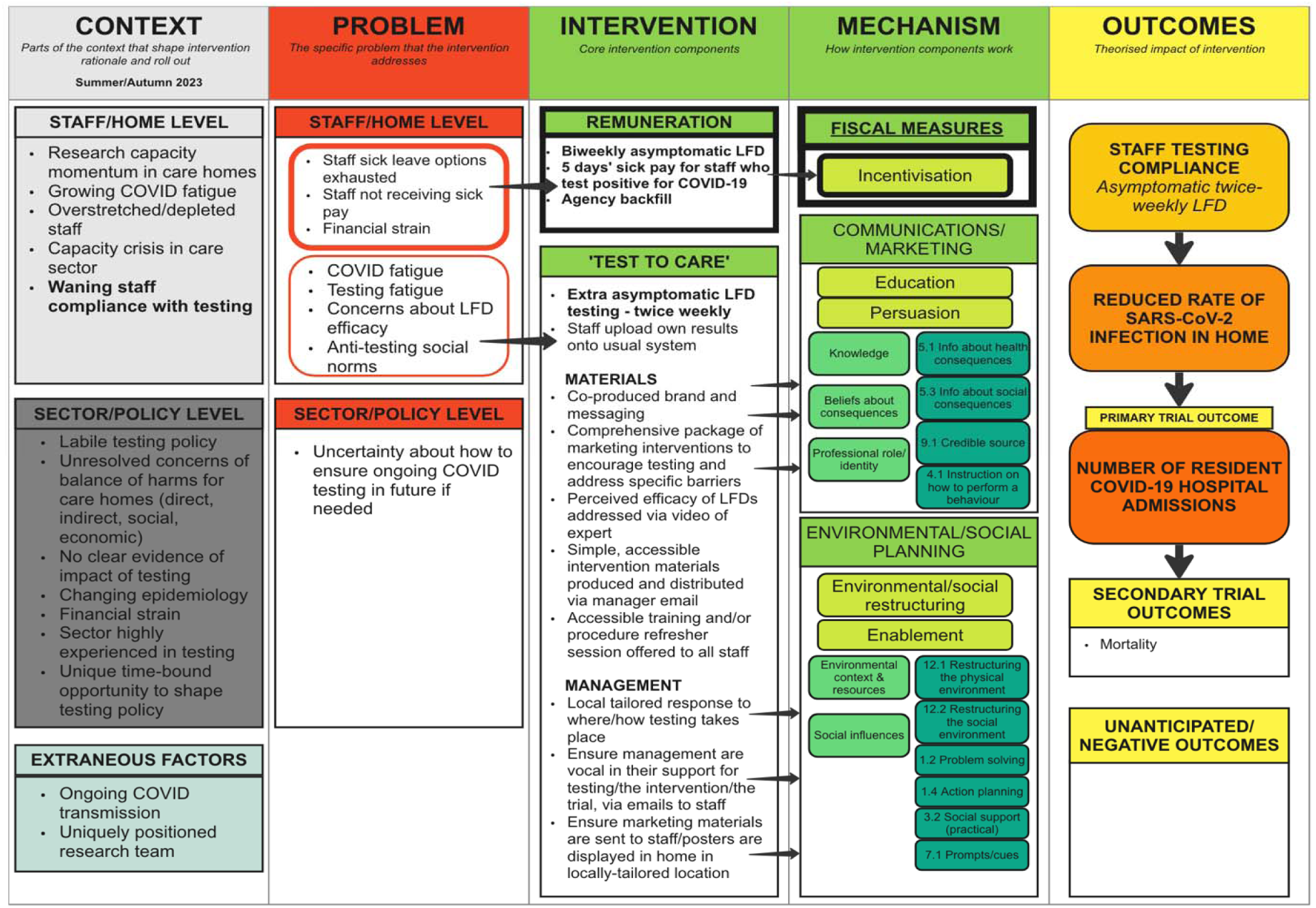
Detailed logic model developed with interdisciplinary feedback.

**Figure 5.**
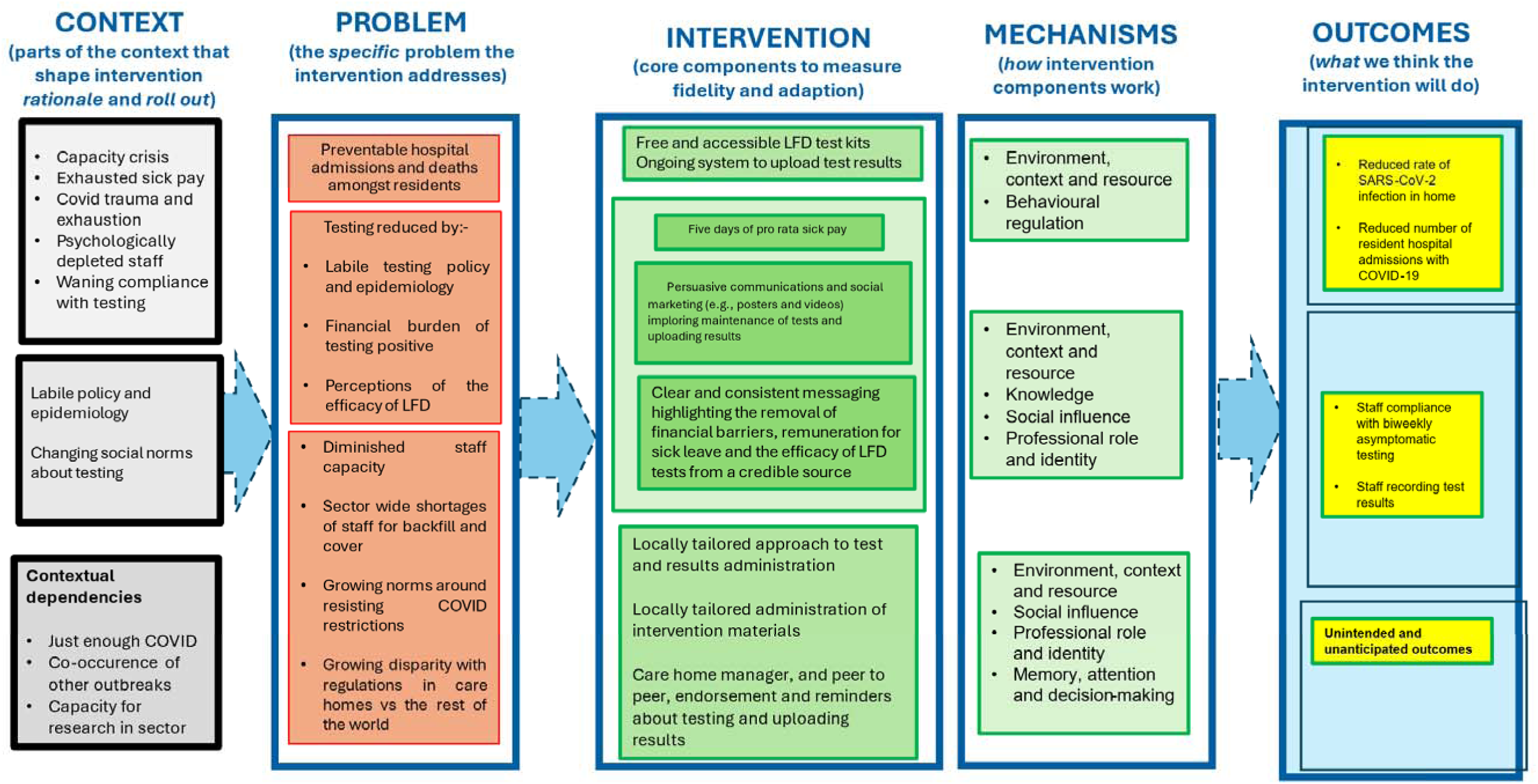
The final logic model developed to map closely to the narrative programme theory (Supplementary file 5)

Figure 3 shows the initial logic model. Figure 4 shows the logic model developed through Phase 3. Finally Figure 5 shows the final pretrial version of the logic model that matches the narrative programme theory.

For Figure 4 RL and PF developed an initial logic model attempting to synthesise and depict *‘Test to Care’*. This initial logic model was circulated to the wider interdisciplinary team for critique and comment. Colleagues’ feedback focussed on ways of improving the depiction of both context and way the hierarchy of outcomes could be described. Minor revisions were made. The revised logic model was then discussed in a series of one-to-one meetings with public and patient representatives. Overall, they found the logic model to be a useful way of depicting the intervention but the idea of a logic model and its reliance on visualisation was a challenge for some. Subsequently, the logic model written out for people who found it difficult to follow the graphic version to also illustrate the underlying programme theory. This full narrative programme theory of *‘Test to Care’* including versions of the intervention material is available as supplementary file D and will shape the planned process evaluation. On its completion a further version of the logic model was completed (Figure 5).

## Discussion

This study aimed to develop an intervention to promote the maintenance of biweekly care home staff self-testing for asymptomatic SARS-CoV-2 with LFDs to be used within ‘VIVALDI-CT’ (Adams et al 2023), a multisite randomised control trial. We show how, within a rapid time frame (around six months), insights from diverse inputs were cumulatively synthesised to develop a scalable, low-intensity intervention for a large clinical trial. Here, we provide a novel exemplar of a four-phase intervention development process within a single study, aligned with leading contemporary intervention development guidance (Skivington et al., 2021; O’Cathain et al., 2019). While other examples of intervention development exist, compared to this, they tend to be partial and not fully aligned to current guidance, for example, use only the BCW approach (e.g., Barker et al, 2016; Loft et al., 2017; Munir et al., 2018, Webb et al., 2016). Additionally, only a small minority of these intervention studies use logic models to describe their interventions (Barker et al., 2018; Murphy et al., 2023) and none provide explicit programme theory explaining what is depicted within their logic models (i.e., supplementary file D) and as a result there is a lack of intervention specification. Therefore, here, we make an important contribution to the intervention development field by outlining a comprehensive, transparent example of intervention development, reporting multiple analyses and processes within a single place. Equally, the main findings discussed in a phase-by-phase overview below contribute novel findings regarding barriers and facilitators to staff testing in UK care homes and specific intervention content to overcome the barriers and enhance the facilitators.

In Phase 1, we conducted a rapid systematic review of published literature then extracted and synthesized available insights into barriers and facilitators to testing. To our knowledge, this represents the best source of data available that addresses the international literature in this field. Barriers and facilitators were multi-levelled (e.g., the sector, the home, the individual care home staff member) and reflected the brief history of the COVID pandemic and responses to it (for example, initial struggles to acquire test kits as availability was limited). Using the BCW approach (Michie et al., 2014), we then developed evidence-based and theoretically-informed intervention content that addressed the barriers and enhanced the facilitators (for example: remuneration for sick leave following positive results; agency staff backfill to cover associated staff absences; communications and marketing persuading staff to keep testing because of the consequences for peers and residents). This phase represents a normative approach to intervention development, using the BCW (e.g., Barker et al, 2016; Loft et al., 2017; Munir et al., 2018, Webb et al., 2016).

In Phase 2, we used a series of stakeholder events to co-produce actual intervention content from the ideas suggested by Phase 1. Little guidance, and few examples, are available which show *how best* to conduct stakeholder engagement as part of intervention development. However, one scoping review exists on the intervention mapping approach and charts the available literature in that field (Bartholomew et al., 1998). Equally Byrne (2019) provides an overview of relevant stakeholder issues and notes the lack of guidance. This lack of literature highlights the poor reporting of, and need for more research around, stakeholder engagement (Majid et al., 2018). We hope our narrative account of the process by which, over time, we combined the perspectives of diverse stakeholders with the results of our BCW analysis offers an example of an approach that could be adapted and built upon within other projects. Actual intervention content, building from the BCW analysis, was progressively developed across a series of events with a range of diverse stakeholders (e.g., front line staff, managers, academic experts). These focussed on rejecting some of the initial intervention ideas (e.g., local testing champions), developing new ideas fit for the actual context in which the intervention would be delivered (e.g., local solutions to the location of testing), and the further specification of the main intervention components, main messages, brand, and modes of delivery in granular detail.

In Phase 3, we conducted a confirmatory qualitative study exploring the ongoing relevance of the barriers and facilitators identified in Phase 1 and shaping Phase 2. We were particularly interested in identifying any novel barriers and facilitators, and considering whether these suggested the intervention needed further modification. Although a few new issues were found (e.g., care home sector exceptionalism in relation to COVID restrictions), none required major changes to the planned intervention. This particular step, in this particular study, related to the fast pace of change in the global COVID-19 pandemic and the labile context of the epidemiology and impact of COVID infection. For other studies set in less labile contexts, or where the published evidence stems from contexts similar to that in which the planned intervention will work, this step may not be necessary.

In Phase 4, we articulated our intervention’s programme theory through both narrative (see supplementary file D) and visual formats (the intervention logic model to visualise the programme theory, Figure 3). We achieved consensus in this process through dialogue with peers and stakeholders (e.g., PPIE representatives) to generate the intervention materials.

As a whole, the intervention development study reported here has shown that it is possible to co-produce a theoretically informed and evidence-based intervention within the English care home sector within a compressed time frame. It is increasingly acknowledged that it is challenging to deliver intervention research in this particular sector and that co-production and stakeholder engagement are essential for intervention traction (Preyer et al., 2022). It is critical that we develop momentum and skill in developing and evaluating interventions within this setting as the need for them will only increase in a world facing the effects of climate induced extreme heat or wider infection prevention and control interventions for example. This study has provided a transparent exemplar of one approach to intervention development. However, we do not claim that it is the sole approach that should be used by health psychologists and other behavioural scientists across diverse behaviours, settings, populations or issues. It is highly likely that diversity in intervention development will enrich the field and lead to maturity in methods and approaches.

### Strengths and limitations

The strengths of the study include the novelty of the approach and our transparent account of how the intervention was developed across a compressed time frame. Such detailed accounts are lacking and their provision will help improve intervention development within and beyond health psychology. Timely intervention development is particularly useful in the context of infectious disease and pandemic infections in particular. Our use and combination of multiple complementary current frameworks and guidance (e.g., the Index study, the BCW approach) to develop the intervention was unique to this study and ensured it was comprehensive and robust. We drew upon each framework proportionately and combined the strengths of each approach. We believe this to be a particular strength of the paper.

A further strength was the use of programme theory and accompanying logic model to provide both a granular and high-level account of the intervention, how it should work, the context, and specific expected outcomes. This has led us to generate a complex intervention with clear discernible interacting parts which have been theorised. Furthermore, we are clear about the logic by which the intervention is intended to work. In addition, our inclusion of multiple stakeholders and PPIE in the development of the intervention allowed for varied perspectives from diverse experts to be a central part of the intervention development process. This included close collaboration with policy makers to ensure that we designed a pragmatic intervention that could be implemented and scaled.

A limitation of the study was our overreliance on qualitative methods alone, and the small sample sizes we were able to recruit within some phases (e.g., Phase 3), potentially limiting the generalisability of the findings to wider populations. A further limitation was the order in which we wrote the programme theory and the logic model: given the time pressures, we drafted the logic model first to use to communicate the intervention ideas with various stakeholders, and drafted the programme theory after. On reflection, the written narrative programme theory may have been more accessible to stakeholders than graphic representation alone (i.e., the logic model).

### Implications

As this paper presents an intervention development study, the success of the ‘Test to Care’ intervention remains unknown. The programme theory will be used to structure a retrospective process evaluation of the intervention. However, given that the current paper has a strong methodological focus and provides an exemplar of a multi-phased approach to intervention development, it may be very useful to health psychologists and others interested in methodological development. However, there are several ways in which the study design used here could be optimised in other studies. These include more expansive and mixed methods for both Phase 2 and Phase 3 which could combine and integrate the best of both quantitative and qualitative research. Equally, Phase 2 could have been enhanced with formal consensus building approaches such as modified Delphi, and the inclusion of designers and visual communicators to complement the health psychology.

## Conclusion

We provide a novel and transparent example of how to develop an intervention using a range of inputs (e.g., published literature, diverse stakeholders (care charities, front line workers, interdisciplinary experts, policy makers) and multiple complementary conceptual and applied frameworks (e.g., MRC guidance, the INDEX study, the BCW, Programme theory and the SEM) to triangulate multiple perspectives and develop comprehensive theory-and-evidence based intervention content. Here, we identified barriers and facilitators to testing and co-produced a practical and scalable intervention for biweekly staff COVID-19 testing in English care homes that will be trialled and evaluated through a qualitative process evaluation. The resulting intervention is a low intensity, scalable intervention that relies on a combination of fiscal measures (financial incentives), communication and marketing and enables change by modifying the environment in which care home staff work. Moreover, as the reported intervention development process was conducted in a compressed time frame, there are useful lessons learned that could be of benefit to others (e.g., using qualitative and quantitative processes within the stakeholder engagement, or developing programme theory from each input to combine at a later date).

## Supporting information

Programme theory

## Data Availability

Some aspects of data are available upon reasonable request (scoping review and BCW analysis). Interview participants did not consent to data sharing.

## Acknowledgements

We would like to acknowledge the support from all the care homes that took part in this project and gave their valuable support and expertise. We would also like to acknowledge the support of the independent members of the Joint Trial Steering Committee and Data Monitoring Committee (TSC-DMC)

