## Supplementary material for "Developing an intervention to maintain biweekly asymptomatic SARS-CoV-2 testing amongst English care home staff: An integrative approach": Programme theory

**Supplementary file: ‘*Test to care*’: Initial Programme theory**

*How we operationalise our Programme theory and depict it in our Logic model*

In this supplementary file, we provide a narrative account that describes how the intervention ‘*Test to Care’* is intended to work. To do so we use programme theory (Skivington et al., 2021) and theorise how the intervention is anticipated to work within a particular context. Specifically, we draw on ideas stemming from realist evaluation and the ‘context, mechanism, outcome’ configuration (Pawson and Tilley, 1997). In the narrative programme theory that follows we explain, in turn, the interactions between 1) The *context* in which the intervention is implemented; 2) The *problem* the intervention is designed to address; 3) The *components of the intervention* itself; 4) The *mechanisms* by which the intervention is intended to work; and 5) The intended and potential unintended *outcomes* of the intervention. In the sections below, we explain the importance of these five particular areas. The narrative account that follows should be read alongside our high-level visualisation - the logic model (Figure 2 within the paper and S1below).

*Figure S1 The final logic model illustrating ‘Test to Care’ programme theory*

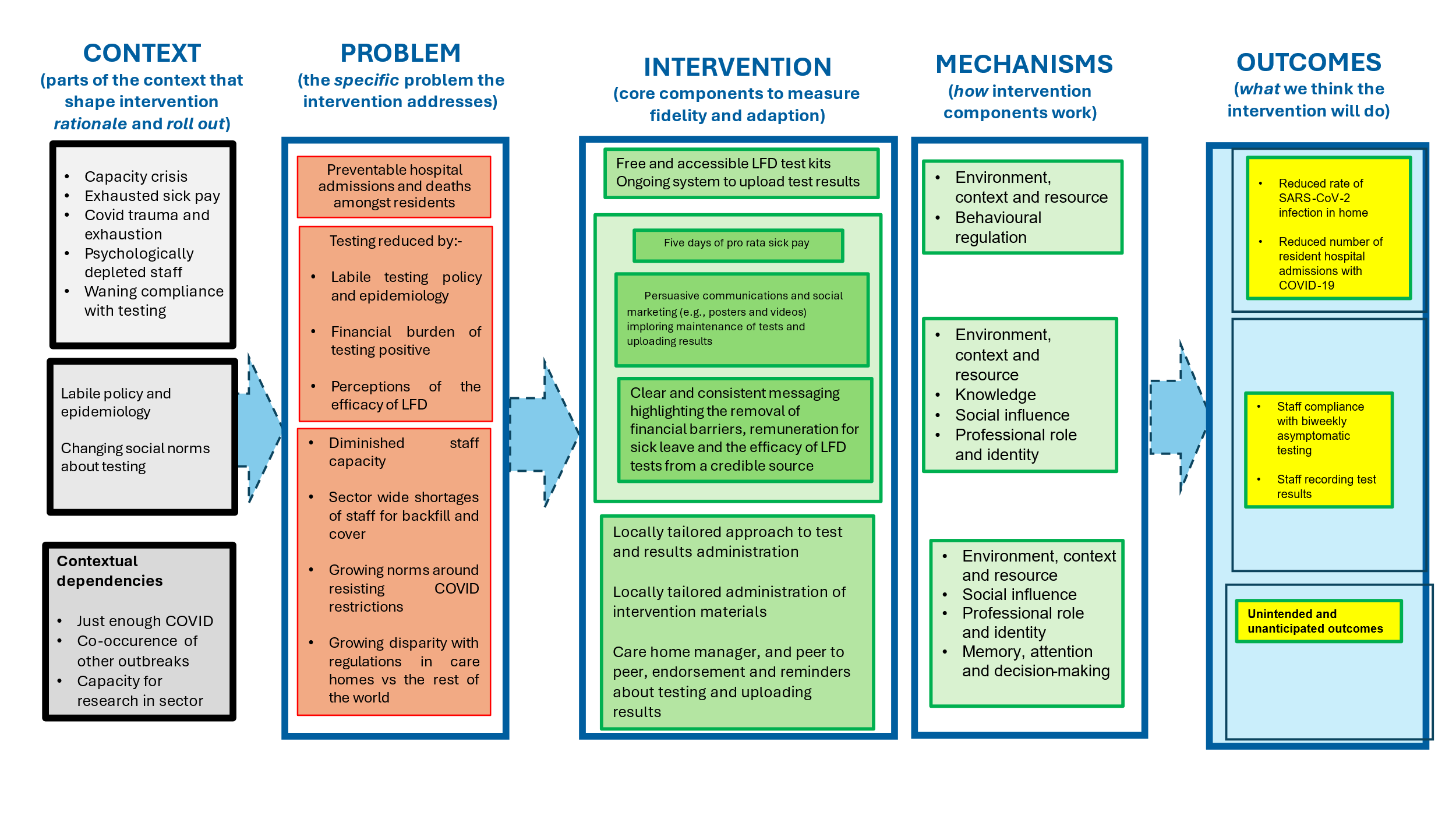

1. *The* ***context*** *(elements of the context shaping intervention rationale and roll out)*

*Why should programme theory detail context*? It is widely acknowledged that context is important in understanding how and why interventions work, for example, see Cambon et al. (2019), Craig et al. (2018), Minary et al., (2018), Moore et al. (2017), Murray et al. (2010) and Shoveller et al (2016). Describing the context in which an intervention is intended to work in detail is important given the limits of intervention transferability across diverse contexts (e.g., Cambon et al., 2012). Equally, describing the important elements of the context that enable or constrain the intervention working as anticipated is vitally important (i.e., the intervention’s ‘contextual dependencies’). An overview of the context is provided in the left-hand column of Figure S1. A more detailed description of Context is provided in Figure S2 below.

*The care home setting*: The care home sector has been uniquely burdened with the most severe negative impacts of COVID-19 given the biological vulnerability of many residents, the overstretched and depleted workforce, and heterogeneity in access to local public health and clinical support due to fragmentation of the care sector. Care home staff remain relatively underpaid compared to other health and social care professionals and the intervention was delivered at a time of wider financial strain within the UK (i.e., the ‘cost of living crisis’, Broadbent et al., 2023). The sector has experienced the highest number of COVID-related deaths (Gordon et al., 2020; Levin et al., 2022) and the greatest degree of restrictions, compared to other health and social care settings (e.g., hospitals) and these restrictions have continued far longer (Hofschulte-Beck et al., 2021). Many care home staff have experienced major trauma (e.g., the impact of COVID-related illness and death in both residents and colleagues, see Gray et al., 2022). Our stakeholder engagement work (Phase 2 of the intervention development process, see this paper) highlighted a sense of growing disquiet in relation to the care home sector and its staff being regulated far more than in other settings (e.g., a condition of ongoing employment was to be fully vaccinated against COVID). We also noted deep heterogeneity across the sector, with some homes running as family-like units, with some staff having worked at the specific care home for many years with long established and close relationships with colleagues, residents and their families. In contrast, other homes were characterised by constantly changing staff members and teams. In some of these latter homes particularly, but not exclusively, high levels of COVID fatigue were reported alongside a waning compliance with COVID-related restrictions and growing resistance to being constantly ‘told what to do’ without consultation. There was also a sense that the particular contribution of the care home sector in addressing COVID-19 had not fully been recognised by comparison to healthcare workers (e.g., clapping for carers was often understood as clapping for the NHS, Manthorpe et al., 2022) and that carers were often asked to comply with restrictions and guidance through processes described to use as akin to ‘emotional blackmail’.

In addition, in some areas of the sector, there is a lack of sick pay and a high prevalence of zero-hour contracts which can drive COVID transmission (see Patel, 2023). For some staff, sick leave allocations have already been depleted via repeated COVID infections in staff and their families, or through other illnesses. As such, there are major financial barriers to asymptomatic testing. The brief and changing history of testing was also important in the sector. Across the duration of the pandemic, testing had at times been mandatory, and a highly-regulated daily task, staff had been tasked with testing themselves and residents and logging all results on to national data systems. Staff were highly skilled at testing using a variety of testing kits. At differing time points in time, staff had tested using a variety of testing technologies as they were developed (e.g., LFDs and confirmatory PCR testing) and some had become aware of the disparity between different types of test in providing accurate results.

*Labile policy contexts*: In relation to our understanding of the context in which ‘*Test to Care’* is intended to work, it is firstly important to acknowledge the labile nature of the COVID-19 pandemic and the associated dramatic shifts in context. Across the COVID-19 pandemic overall (2019-2023), there were major changes in epidemiological, biological, and psychosocial aspects of COVID-19. Its incidence and virulence changed according to both the evolution of SARS-CoV-2 and population-level immunity because of being vaccinated against, or exposed to, the virus (e.g., see Looi, 2023). Across the brief history of this pandemic, COVID-19 related legislation and guidance was also markedly fluid, alongside changing social norms and individual beliefs (e.g., about mask use, or vaccination). It is this ever-changing policy context that shaped the need and funding for the wider study (‘VIVALDI-Clinical Trial’, see Adams et al., 2023) in which this intervention development study is embedded. Therein we hoped to address the key policy question of whether asymptomatic testing is effective and cost-effective. The design of our intervention development study, with its use of diverse inputs across time, attempted to ensure the intervention was fit for purpose for the particular temporal and cultural context in which it was delivered.

*Figure S2. Details relating to Context that figure within the logic model*

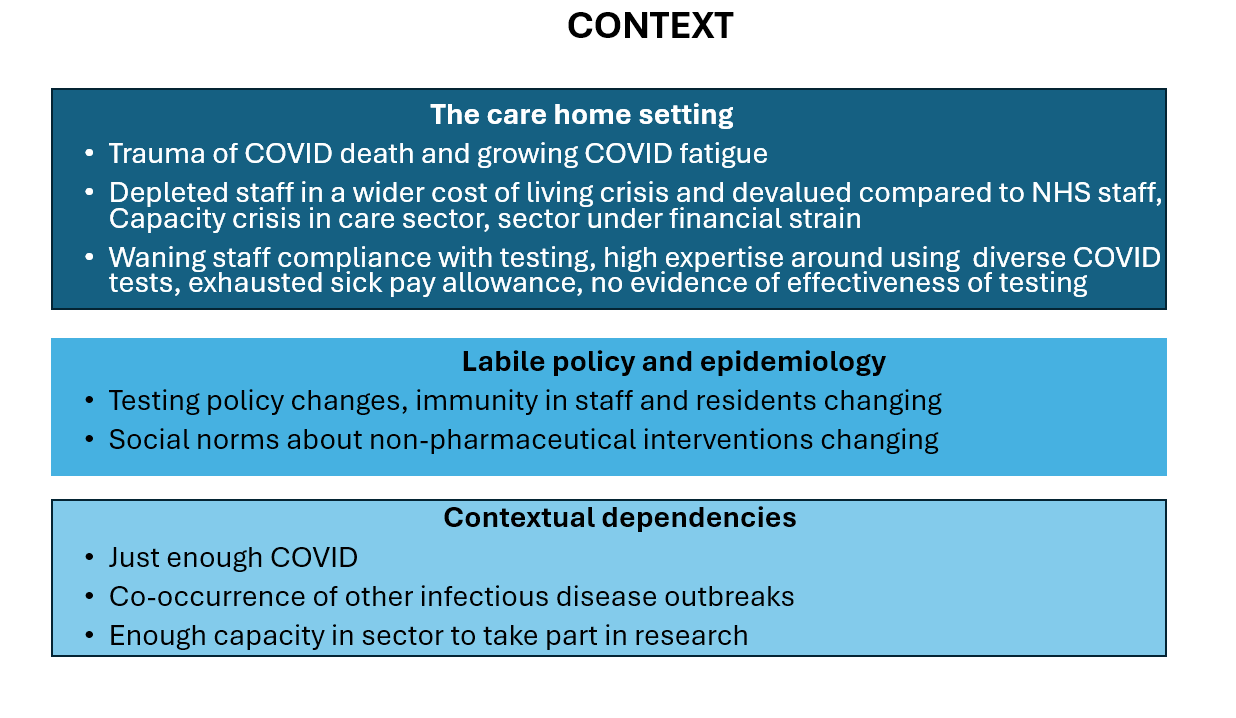

*Contextual dependencies (anticipated factors critical to the intervention having traction)*: We predicted that for a testing intervention to work, there must be ‘just enough COVID’. In other words, there is a ‘Goldilocks zone’ wherein the intervention may gain traction and prove effective, but we can also anticipate circumstances in which this will not be possible. At one extreme, if there is ‘too much’ COVID infection, then individual care homes and the sector overall risk being overwhelmed, and the intervention compromised. Diminishing capacity to test, cover staff sick leave, and avoid outbreaks closing the home, may halt any meaningful implementation of the intervention and represent a major barrier to the intervention working as anticipated. At the other extreme, it is also essential that ongoing COVID transmission occurs and that COVID infection continues to lead to hospital admissions (i.e., the primary trial outcome). Furthermore, it is also vital that national testing policy (i.e., the control arm of a future trial) remains distinct from the intervention arm (i.e., ‘*Test to Care*’). Other important contextual dependencies include the co-occurrence of other infectious diseases (e.g., pandemic influenza) and that the sector has enough capacity to participate within a trial and provide sufficient high-quality on-going data to enable meaningful analysis of the trial (i.e., VIVALDI-CT) and its effectiveness.

2. The **problem** (the specific problem(s) that the intervention is designed to address)

*Why should programme theory detail the problem addressed by the intervention?* It is essential that those developing and specifying interventions clarify the precise problem that they consider the intervention resolving. ‘*Test to Care’* is intended to address the multi-levelled biopsychosocial (Engel, 1977) problems associated with COVID-19 within the UK care home context in 2023. Whilst many of these have been touched upon within the context section above, here we use the biopsychosocial framework to structure and specify the precise problems the intervention is intended to address. An overview of the problem is presented in column 2 of the logic model but Figure S3 provides more detail below.

*Figure S3. Details relating to the problem that figure within the logic model*

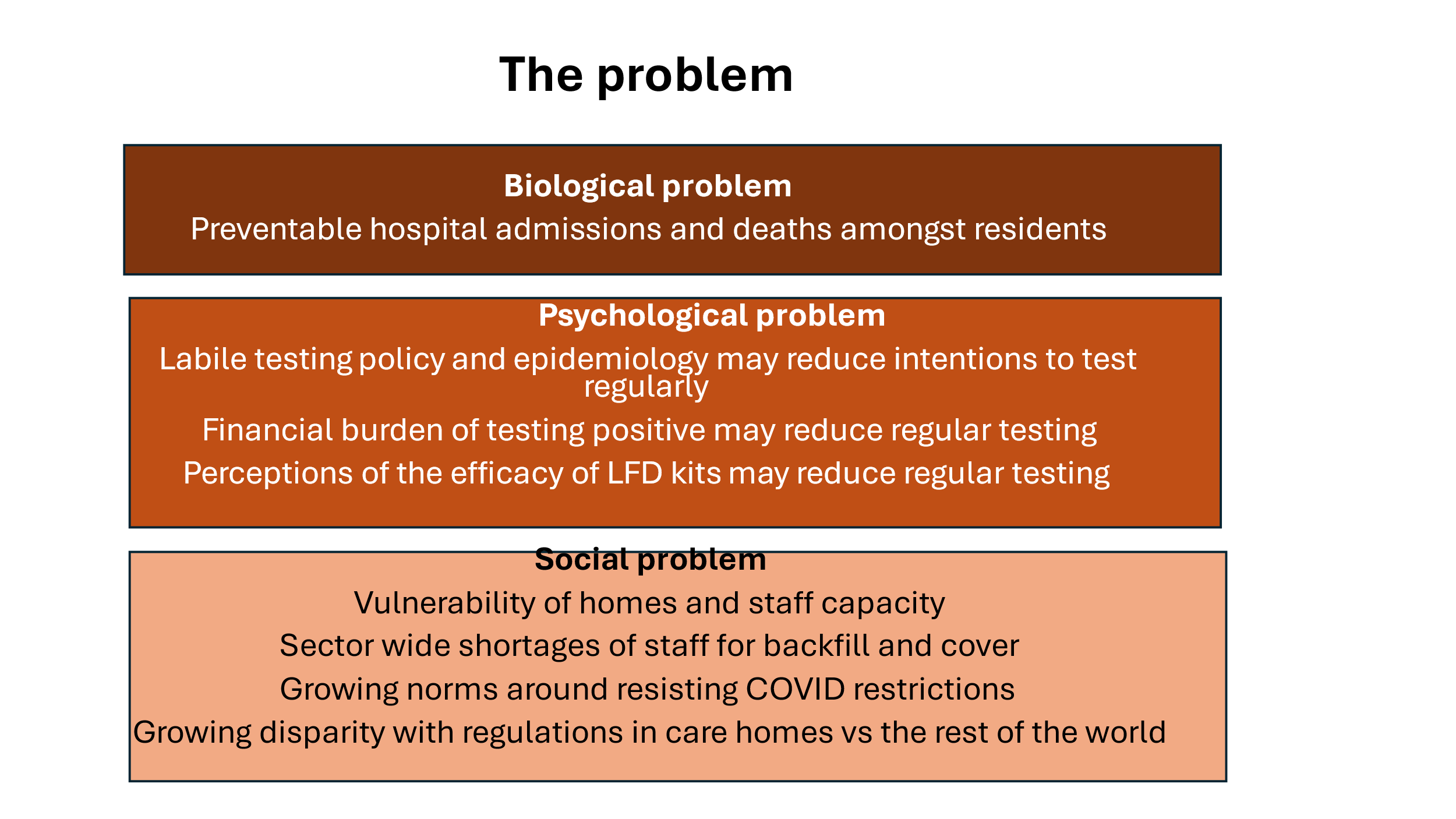

*The biological problem*: The intervention must reduce both the number of care home resident hospital admissions and deaths associated with SARS-CoV-2. Undiagnosed SARS-CoV-2 infection amongst care home staff is assumed to drive resident admissions and deaths, although the proportion of infections resulting in severe outcomes will vary depending on the pandemic stage / SARS-CoV-2 variant in circulation.

*The psychological problem*: For care home staff, decisions to maintain asymptomatic testing for SARS-CoV-2 in 2023 are not straightforward (see Table 3 in main paper). Testing is no longer mandated by government or employers. Anxiety and worry over potential positive test results was commonly reported in the intervention development work. Critically, the financial risks associated with testing are very important aspects of these concerns; testing positive with asymptomatic infection could result in the loss of income at a time of national financial crisis. Finally, staff concerns relating to the perceived efficacy of lateral flow device (LFD) test kits is a further barrier to testing. Echoing this, it is vital that staff perceive the link between their testing and residents’ health outcomes (e.g. if staff see that residents get infected but remain well the argument for testing is diminished). Other complex barriers to maintaining testing include growing levels of COVID fatigue, and a strong desire to put the trauma of the COVID pandemic into the past and ‘get back to normal’. In contrast to these barriers about testing itself, acquiring the skills to test correctly, and normalising testing are not major problems in relation to maintain testing (given the history of testing throughout the COVID pandemic and the intervention does not have to teach people how to test. However, the intervention must focus on highlighting the need to focus on *maintaining* testing over time rather than the *adoption* of testing *per se* (see Kwasnicka et al., 2016 for more on the importance of maintaining behaviour change over time).

*The social problem*: The intervention must address the particular vulnerability of care homes themselves, as staff capacity to maintain care home function is fragile and could be compromised by large numbers of staff testing receiving positive results and being off sick. The intervention must ensure that staff who test positive are enabled to stay at home and avoid transmission occurring within the workplace. This minimises the chance of residents being infected and needing hospital admission. Moreover, for the care home sector, there is a risk that struggling to replace staff who receive positive results may add to the financial burden of care homes and diminish care home function. Other socially situated problems include growing social norms that resist further COVID restrictions and seek to harmonise the regulation of care home staff with the rest of society (e.g., removal of masks, the cessation of testing).

3. The **intervention** (its core components)

*Why should programme theory detail the components of an intervention?* The importance of describing intervention components is well-known (see Clark, 2013; Hoffmann et al., 2014; Michie, Atkins and West, 2014; Sutcliffe et al., 2015). ‘*Test to Care’* must be a complex intervention with multi-levelled interacting parts reinforcing each other and directly addressing the multi-levelled biopsychosocial problems articulated in the ‘problem’ section above. It is vital that the intervention being trialled is scalable and sustainable, minimising any further burdens on care home staff, care homes themselves, and the care home sector. The central column of the logic model provides an overview of the intervention components, but further visual detail is shown below in S4. Figure S4 also uses the ‘intervention functions’ and ‘behaviour change techniques – BCTs- from the BCW approach to describe the intervention components.

*Biomedical components*: To reduce the incidence of SARS-CoV-2 within care homes and associated resident hospital admissions and deaths, biweekly staff testing to identify asymptomatic infection must be enabled through the simple, ongoing, provision of free and accessible LFD test kits. These were supplied for the purpose of the trial by the UK’s Health and Security Agency (UKHSA). Equally, it is important that the same systems to upload and record the results of tests used throughout the COVID-19 pandemic are maintained and available to staff to minimise any reporting burden. Given the desire for future scalability and likely cost-effectiveness the use of confirmatory PCR testing to confirm LFD test kits results is not necessary or advised. These intervention components relate to the BCW’s intervention functions (Michie et al., 2014) ‘environmental restructuring’ and ‘enablement’ and can be further operationalised using Behaviour Change Techniques (BCTs) from the BCT Taxonomy version 1 (BCTTv1; Michie et al., 2013), ‘2.6. Biofeedback’ and ’12.5 Adding objects to the environment’.

*Figure S4. Details relating to the intervention components that figure within the logic model*

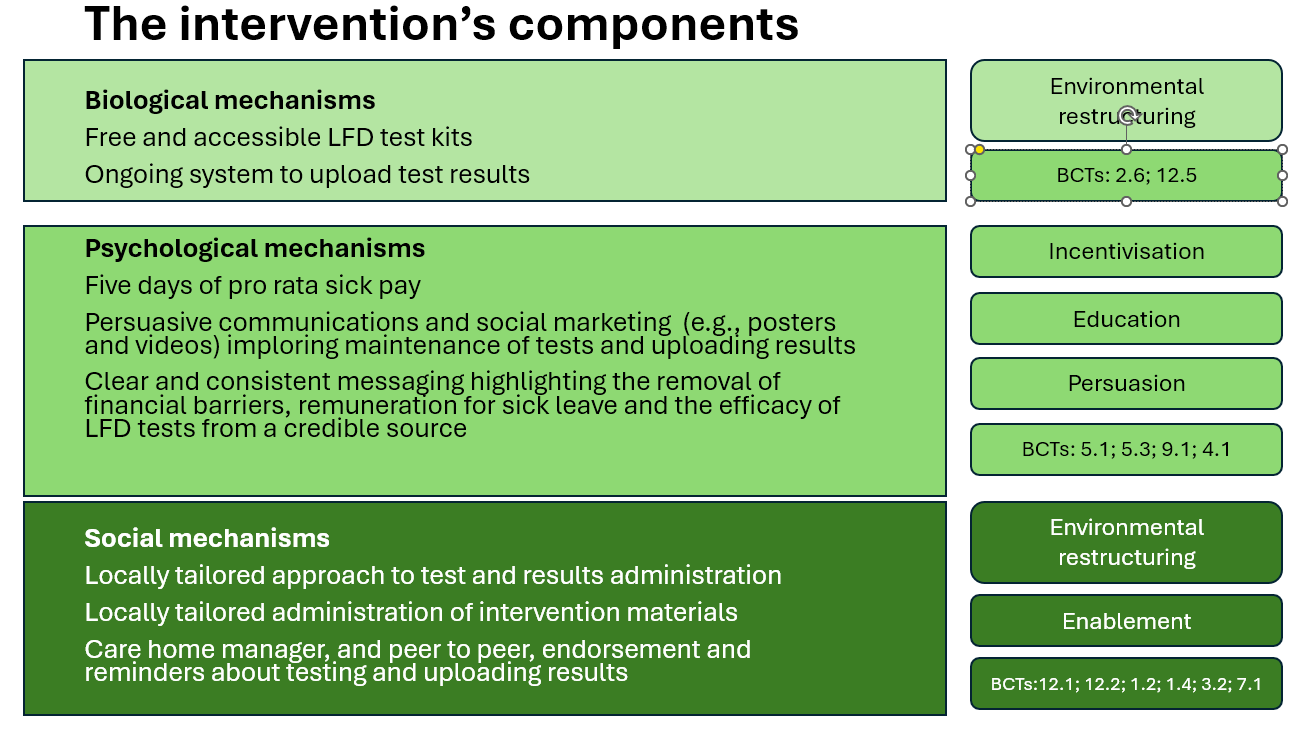

*Psychological components:* To address key barriers to testing and reduce worries over test results, the intervention should, first and foremost, incentivise regular testing through the removal of financial concerns around sick pay. As such, the intervention included five days of pro-rata sick pay provided by UKHSA. Accessible financial incentives to keep testing should also be clearly communicated to staff consistently. We anticipate that, amongst homes with varied beliefs about the ongoing value of testing (e.g., where some people want to test and others do not), the fact that some staff who test positive will receive sick pay following regular asymptomatic testing, should provide a positive feedback loop over time, modelling the value and importance of regular testing and setting positive norms about testing within given care homes.

In addition to providing simple clear mechanisms to access sick pay, complementary components should revolve around the use of communications and social marketing (e.g., posters and videos, see Figures S1 and S2) to persuade staff to maintain testing and to remind them to upload test results over the duration of the intervention.

In relation to clear and consistent messaging highlighting the removal of financial barriers to testing, the possibility of remuneration should be made clear: ‘*Test with peace of mind knowing you will get sick pay if you test positive*’. Critically, these communications should also address barriers to testing that focus on beliefs around the efficacy of LFD kits to provide accurate test results. As such, communications should include a credible source affirming the value and efficacy of LFD kits for their identification of SARS-CoV-2. We chose to operationalise these messages with a brief video of a senior health and social care professional talking persuasively about how the kits were ‘good enough’ for the study. A link to this video was included within an email to staff sent from their care home manager (<https://tinyurl.com/jcy4ucds>).

Given the normative and habitual nature of testing within the sector, only simple and consistent messaging around what carers are being asked to do is required within the intervention: ‘*Test to Care’:* *do extra testing, twice a week, because you care*’, wherein ‘extra’ refers to the asymptomatic and ongoing nature of the testing. Owing to sensitivities within the sector, the intervention and its materials and branding should draw upon, but not focus entirely, on the professional role and identity of carers. Our stakeholders had suggested the branding ‘*Test to Care’* to highlight the connection between carers’ roles and testing and, drawing on the familial nature of many care homes, to focus on the altruistic aspects of caring ‘*test to protect each other – staff and residents’* rather than individual health benefits (in a context where vaccination and immunity through exposure was common). As staff already have expertise and skills around testing, and the uploading of results has been usual practice across the COVID-19 pandemic, a light-touch approach to further training and reminders to test and upload test results is considered appropriate. The intervention should also use familiar and existing resources already branded as credible sources. Signposting to these resources is appropriate and is able to accommodate new carers joining the sector, in addition to accommodating any changes to the content of LDF kits. To avoid telling staff ‘what to do’, the intervention will suggest ‘*if you want to refresh your COVID testing techniques, see’* etc).

***Figure S5 An example of the ‘Test to Care’ poster to be displayed near clocking-in machine***

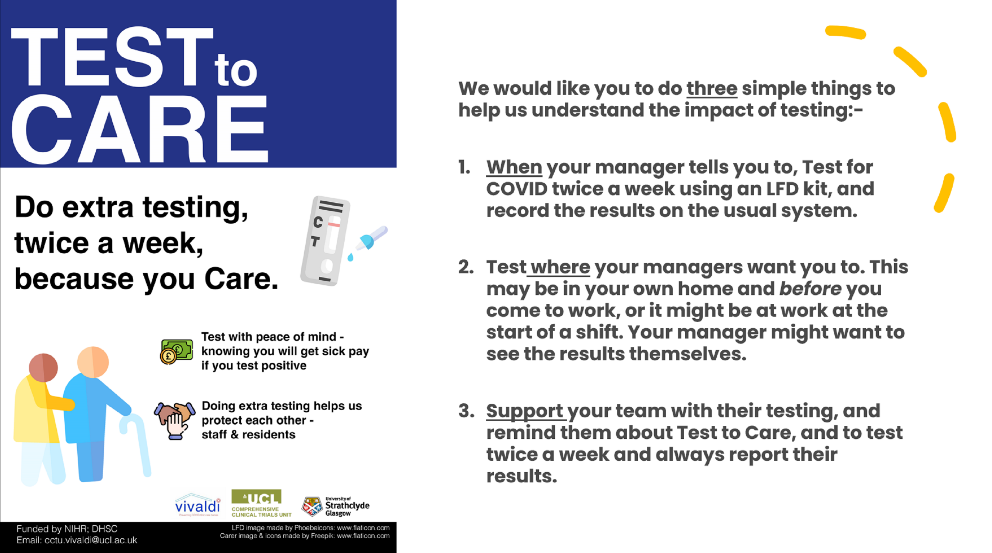

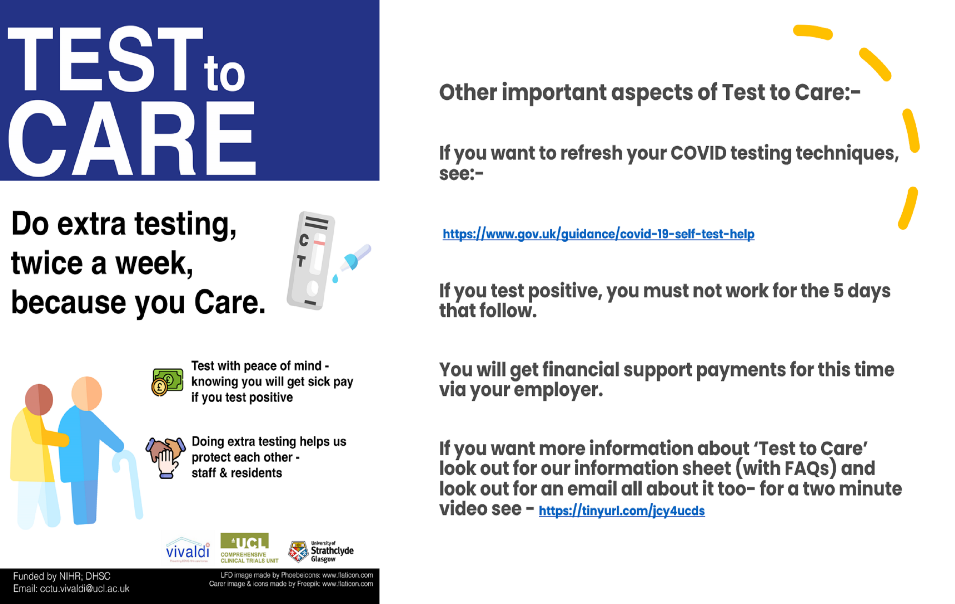

Together these intervention components relate to range of corresponding intervention functions using the BCW approach (Michie et al., 2014) are also employed with the psychological components of ‘*Test to Care’*, including ‘incentivisation’ (i.e., sick pay), ‘environmental restructuring’ and ‘enablement’ (i.e., provision of free LFD kits, local arrangements for testing and recording results), ‘persuasion’ (i.e., manager emails to staff, video of expert affirming efficacy of LFDs, posters), and ‘education’ (i.e., posters and video). These intervention functions can be operationalised further and include BCTs (Michie et al., 2013) such as ‘5.1 information about health consequences’, ‘5.3 information about the social consequences’, ‘9.1 credible source’ and ‘4.1 instructions on how to perform a behaviour’.

*Social components*: Socially-focussed intervention components will complement the psychological ones highlighted above. The intervention will harness social influence and the local social environment in several ways, including enabling a locally-tailored approach to the administration of the processes that surrounded staff testing. For example, care homes could decide themselves where and when staff should test, and how and by whom test results would be uploaded. Managers are also encouraged to decide exactly where to display the intervention processes to best effect (we suggested next to the clocking-in machine). In this way, administrative workload and the imposition of potentially burdensome intervention processes could be avoided and, indeed, local ownership encouraged. Wherever possible, the intervention should capitalise on the cascading of intervention communications, messages, and materials within local managerial and operational structures. Within care home manager communications, and interactions between peers, there should be vocal support in for ‘*Test to Care’* and communications to staff should highlight the importance of testing and remembering to upload test results (managers are also asked to circulate the video depicted outlined above).

These materials are also important in addressing the heterogeneity of beliefs about testing within a given home. Staff with positive results will receive sick pay following regular asymptomatic testing, whereas those who may choose not to test or to upload their results do not. This should provide a salient model of the value of regular testing and set positive norms about testing within given care homes. Increasing numbers of staff testing and getting paid for sick leave associated with reactive results should provide some critical mass within a given care home and visible pro-testing social norms.

Together these components relate to the intervention functions (Michie et al., 2014) of ‘environmental restructuring’ and ‘social influence’ which can be operationalised further by BCTs (Michie et al., 2013) such as ’12.1 Restructuring the physical environment’, ’12.2 Restructuring the social environment’, ‘1.2. Problem Solving’, ‘1.4 Action planning’, ‘Social support (practical)’ (i.e., receipt of sick pay), and ‘7.1 prompts and clues’ (i.e., the placement of the posters).

4. The **mechanisms** (theorised ways in which the intervention is intended to work)

*Why should programme theory detail the mechanisms of an intervention?* An understanding of the mechanisms by which interventions work is vital for both developing and evaluating how an intervention works within a given context. A variety of frameworks are available to consider how to conceptualise mechanisms. These range from the psychological (e.g., Carey et al. 2023), the social (May et al., 2022), or the systemic (Rutter et al., 2017). Given the multi-levelled components by which ‘*Test to Care’* works, its purposed (i.e., hypothetical) mechanisms are diverse; the way the intervention components work can all be understood by using different theoretical lenses. Again, to assist with understanding these complementary mechanisms, the three elements of the biopsychosocial model are used and here we focus on understanding ‘*Test to Care’* primarily through the TDF approach which is part of the behaviour change wheel approach (Michie et al., 2014). The intervention mechanisms are shown in the fourth column but also below in Figure S5

*Figure S6. Details relating to the intervention mechanisms using the TDF that figure within the logic model*

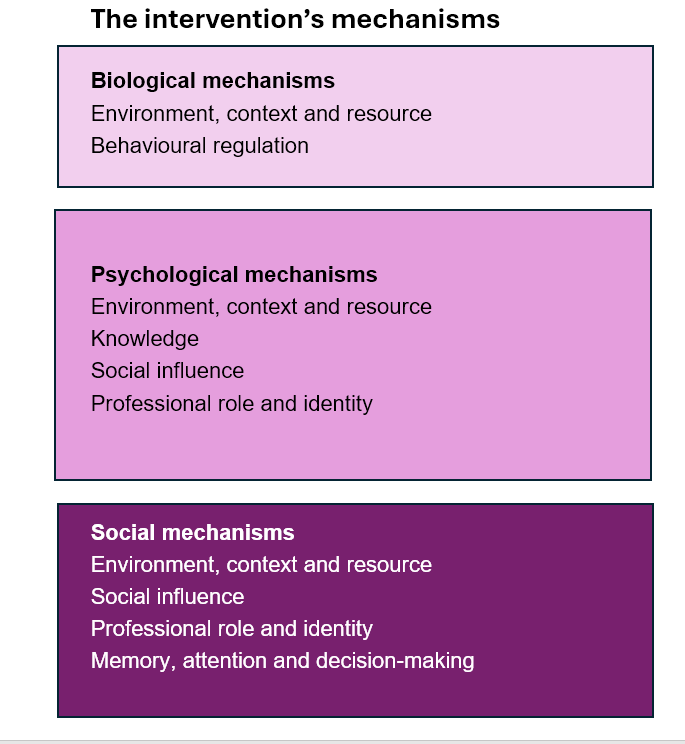

*The mechanisms of the biomedical components*: Using the Theoretical Domains Framework (TDF; Atkins et al., 2017), important theoretical domains used within the intervention include ‘environment context and resource’ via the provision and function of LFD test kits and ‘behavioural regulation’ via the uploading and monitoring of results.

*The mechanisms of the psychological components*: Important TDF domains (Atkins et al., 2017) used within the psychological components of the intervention include ‘knowledge’ (i.e., telling people to do biweekly testing with LFDs, telling people about access to sick pay for positive results), ‘beliefs about consequences’ (i.e., reassuring people about remuneration if they test positive, encouraging people to test to protect each other and residents), ‘professional role and identity’ (i.e., testing because staff care and are in the caring profession), ‘environment, context and resource’ (i.e., ‘the provision of LFD kits, maintaining access to results system, testing at a locally endorsed location‘), and ‘social influence’ (i.e., peers endorsing and encouraging testing, seeing staff taking part and getting paid with positive results).

*The mechanisms of the social components*: Important TDF domains (Atkins et al., 2017) used within the social components of the intervention include ‘environment, context and resource’ (i.e., wherein locally-tailored approaches to operationalising the intervention, such as the location of testing, are developed), ‘social influence’ and ‘memory, attention and decision-making’; (wherein peers remind and encourage each other to test), and ‘professional role and identity’ (whereby key messages and communications are cascaded down local management structures).

The **outcomes** (the theorised impact of the intervention)

*Why should programme theory detail the outcomes of an intervention?* An understanding of the hypothesised outcomes of interventions is central to most research around evaluating complex interventions. Outcome evaluations are commonplace and form the mainstay of traditional trial research. However, there are calls for more nuanced approaches (e.g., Rychetnik et al., 2002) and the idea of programme theory is that it makes clear the connections between context, mechanisms, and outcomes. As such, programme theory should address the relationship between the intervention and a trial’s primary outcome, but also consider a hierarchy of outcomes and include a focus on unintended and unanticipated outcomes. These cannot be specified at an early stage of intervention development, but the programme theory should highlight the need to address them in the future (e.g., within process and outcome evaluations).

*Figure S7. Details relating to the intervention mechanisms using the TDF that figure within the logic model*

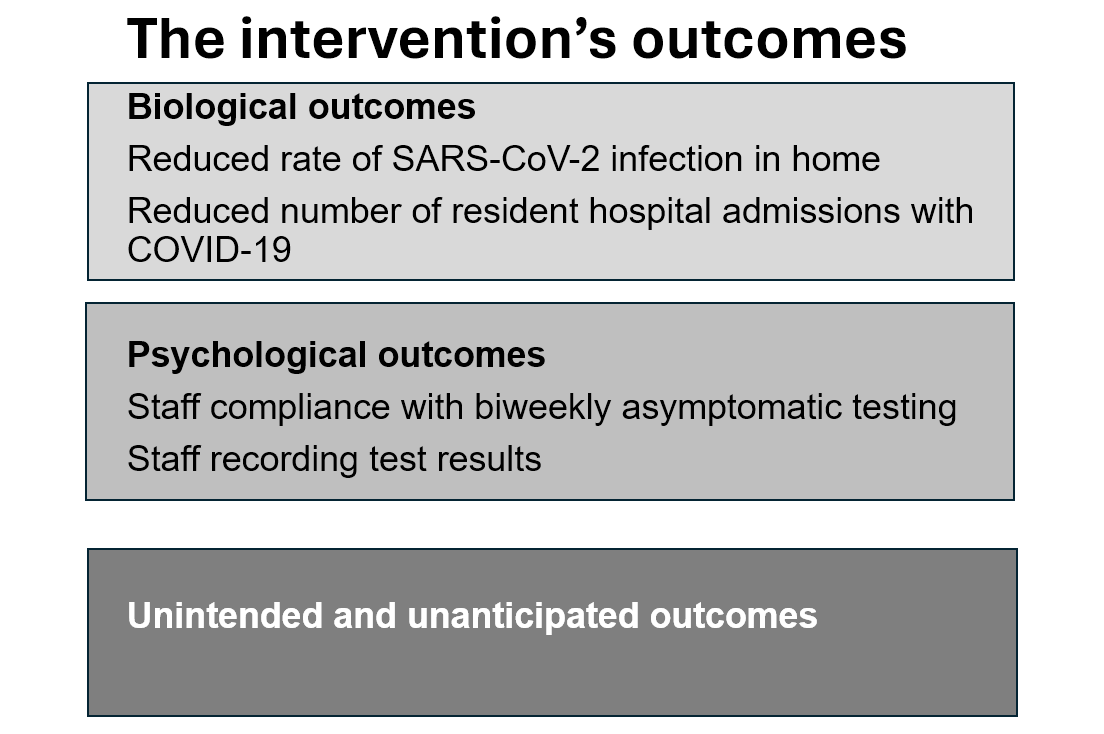

The ‘*Test to Care’* intervention is imagined to change a set of interconnected outcomes. The trials primary outcome is the number of resident COVID-19 hospital admissions. Secondary outcomes, which reflect the interventions central logic include staff compliance with biweekly asymptomatic testing, staff taking sick leave with positive test results, associated reductions in the incidence of SARS-CoV-2 leading to a reduction in resident infections with SARS-CoV-2 and corresponding reductions in COVID-related hospital admissions.

*The necessary fluidity of programme theory*

It should also be noted that the *’Test to Care’* programme theory, unlike a static resource such as an intervention manual, is iterative and expected to change across the duration of the study and beyond. For example, effective interventions change the very context in which they operate, and their programme theory will need to be updated; equally, partially effective interventions need to be optimised by making changes to their content and enhancing implementation. Equally for ineffective interventions, it may be possible to use the programme theory to examine what did not work and why, to direct focussed modifications. Here, given our focus on using programme theory for intervention development, we explicitly describe our programme theory as ‘initial’ to signal its mutability (i.e., the fact that it will change).

Currently, at the time of its development, we already anticipate adaptations and revisions to the ‘*Test to Care’* programme theory in light of insights from the planned process evaluation which will detail what, when, why, and how things work and in which circumstances (Moore et al., 2014). Should the intervention prove successful in relation to the trial’s primary outcome, we anticipate the ‘*Test to Care’* programme theory will provide a useful starting point for transferability of the intervention to other geographic, temporal, and epidemiological contexts. Should the intervention prove ineffective or merely partially effective, the granular detail of our programme theory invites systematic and focussed modification. We also believe our initial ‘*Test to Care’* programme theory could provide a good starting point for considering wider testing interventions for other infectious diseases within the context in which it was developed (for example, UK care homes and testing for influenza).
